# Assessing changes in incubation period, serial interval, and generation time of SARS-CoV-2 variants of concern: a systematic review and meta-analysis

**DOI:** 10.1101/2023.05.19.23290208

**Authors:** Xiangyanyu Xu, Yanpeng Wu, Allisandra G. Kummer, Yuchen Zhao, Zexin Hu, Yan Wang, Hengcong Liu, Marco Ajelli, Hongjie Yu

**Author notes:** Corresponding authors: Marco Ajelli, Hongjie Yu. Joint first authors (Xiangyanyu Xu, Yanpeng Wu, Allisandra G. Kummer). Joint senior authors (Marco Ajelli, Hongjie Yu).

## Abstract

**Background:** After the first COVID-19 wave caused by the ancestral lineage, the pandemic has been fueled from the continuous emergence of new SARS-CoV-2 variants. Understanding key time-to-event periods for each emerging variant of concern is critical as it can provide insights into the future trajectory of the virus and help inform outbreak preparedness and response planning. Here, we aim to examine how the incubation period, serial interval, and generation time have changed from the ancestral SARS-CoV-2 lineage to different variants of concern.

**Methods:** We conducted a systematic review and meta-analysis that synthesized the estimates of incubation period, serial interval, and generation time (both realized and intrinsic) for the ancestral lineage, Alpha, Beta, and Omicron variants of SARS-CoV-2.

**Results:** Our study included 274 records obtained from 147 household studies, contact tracing studies or studies where epidemiological links were known. With each emerging variant, we found a progressive shortening of each of the analyzed key time-to-event periods. Specifically, we found that Omicron had the shortest pooled estimates for the incubation period (3.63 days, 95%CI: 3.25-4.02 days), serial interval (3.19 days, 95%CI: 2.95-3.43 days), and realized generation time (2.96 days, 95%CI: 2.54-3.38 days) whereas the ancestral lineage had the highest pooled estimates for each of them. We also observed shorter pooled estimates for the serial interval compared to the incubation period across the virus lineages. We found considerable heterogeneities (I^2^ > 80%) when pooling the estimates across different virus lineages, indicating potential unmeasured confounding from population factors (e.g., social behavior, deployed interventions).

**Conclusion:** Our study supports the importance of conducting contact tracing and epidemiological investigations to monitor changes in SARS-CoV-2 transmission patterns. Our findings highlight a progressive shortening of the incubation period, serial interval, and generation time, which can lead to epidemics that spread faster, with larger peak incidence, and harder to control. We also consistently found a shorter serial interval than incubation period, suggesting that a key feature of SARS-CoV-2 is the potential for pre-symptomatic transmission. These observations are instrumental to plan for future COVID-19 waves.

## Introduction

As of April 12, 2023, the COVID-19 pandemic has resulted in more than 762 million reported cases and 6.8 million reported deaths [1]. Since the first detection of SARS-CoV-2 (the virus causing COVID-19) in Wuhan, China, in December 2020 the virus has started to evolve, and several variants of SARS-CoV-2 have been identified. Five of them were classified as variants of concern (VOCs): Alpha (B.1.1.7), Beta (B.1.351), Gamma (P.1), Delta (B.1.617.2), and Omicron (B.1.1.529) [2]. Alpha and Delta were particularly successful at spreading around the globe, causing major waves of infections and associated hospitalizations since late 2020 and early 2021[3] [4]. In late 2021, Omicron was first detected in South Africa and spread more rapidly than all other VOCs, causing massive outbreaks worldwide albeit with a lower associated burden due to high vaccination levels [2, 5, 6]. As of March 2023, Omicron is the dominant variant worldwide.

Knowledge of the transmission dynamics of SARS-CoV-2 VOCs is vital for understanding the epidemiology of COVID-19 and establishing effective control measures. In this context, three key indicators are the incubation period, serial interval, and generation time. The incubation period is the interval between an individual’s time of infection and symptom onset [7]; the serial interval is the interval between symptom onset of the infector and symptom onset of the infectee(s) [7]; the generation time is the interval between the infector’s time of infection and the infectee(s)’s time of infection [8]. For example, these three indicators are instrumental for defining the duration of quarantine and which contacts need to be traced as well as providing insights into pre-symptomatic transmission. Moreover, in combination with the reproduction number, they are key for properly interpreting epidemic growth and forecasting epidemic timing [9].

While many epidemiological investigations have estimated the incubation period, serial interval, and generation time for the VOCs, these indicators can be affected by a variety of factors related to the specific epidemiological situation of each study as well as deployed interventions [10-14]. For this reason, it is essential to combine these estimates to provide a more comprehensive picture of these indicators for the different VOCs. However, previous reviews and meta-analyses on the incubation period, serial interval, and generation time for SARS-CoV-2 have not examined the evolution of these three indicators across the different VOCs [13, 15-23]. This information is essential for outbreak preparedness and response planning as SARS-CoV-2 continues to mutate and systematically comparing changes in these indicators can provide insights into their possible future trajectory. Therefore, our objective in the present review is to quantitatively synthesize estimates for the incubation period, serial interval, and generation time and examine how these indicators have evolved over time from the ancestral lineage to the different VOCs.

## Methods

### Search Strategy

We conducted a systematic search following the Preferred Reporting Items for Systematic Reviews and Meta-Analyses (PRISMA) guideline (see PRISMA checklist) [24]. We searched for studies published in English on three peer-reviewed databases (PubMed, Embase, and Web of Science) and five preprint servers (medRxiv, bioRxiv, Europe PMC, SSRN, and arXiv) using predefined search terms (Supplementary Table S1). All searches were conducted on March 28, 2023.

### Inclusion and exclusion criteria

Studies were included if they satisfied the following criteria: (1) provided at least one summarized statistic (e.g., central tendency and dispersion) for the incubation period, serial interval, or generation time; (2) relied on data from a contact tracing study, a household study or a study where epidemiological links were known.

We excluded studies that: (1) were meta-analyses and reviews, study protocols, media news, commentaries, or the full text was unavailable (e.g., conference abstract); (2) SARS-CoV-2 variant/sub-variant or study period not reported; (3) carried out analysis with sample size less than five; (4) were non-human studies; (5) methods were not described.

### Outcome measures

The outcome variables were incubation period, serial interval, and generation time. We further divided the generation time into two categories: intrinsic and realized. The intrinsic generation time represents the generation time that would be observed in a fully connected infinitely large susceptible population in the absence of interventions and behavioral change. The realized generation time refers to the generation time that is observed in field condition and is thus affected by specific features of the study population (e.g., interventions, individual behaviors, analyzed social settings) [17, 25, 26].

### Data extraction

Study screening and data extraction were performed independently by authors YP.W. and X.X., and inconsistencies were reconciled together with a third author, Y.Z. For eligible studies, we extracted all summary statistics related to the outcomes of interest, including mean, median, interquartile range (IQR), range, standard deviation, quantiles, 95% confidence interval (CI), and 95% credible interval (CrI). In addition, we collected descriptive data on the authors, article’s title, journal, date of publication, study location (country or region), date of the study period, total number of study subjects, variant type, and methods used for estimating key time-to-event intervals.

### Quality assessment

Two authors (Z.H. and Y.Z.) independently carried out the quality assessment of the included literature. The quality assessment scale was adapted from the Newcastle-Ottawa quality assessment questions (detailed in the Supplementary Table S2) [16]. Disagreements between the two reviewers were resolved together with a third author, X.X.

### Data analysis

We described the central tendency (mean or median) and dispersion measure (range, IQR, 95%CI, or 95%CrI) for incubation period, serial interval, and generation time for the different VOCs via a forest plot. For studies reporting both mean and median, we preferred the mean estimates since most studies reported mean. For dispersion measures, we preferred standard deviation, followed by 95%CI/CrI, IQR, and range.

Next, we conducted a meta-analysis for different SARS-CoV-2 variants. The random- or fixed-effects models were employed for the pooling analysis according to the heterogeneity test. Notably, if standard deviation was not available but the parametric distribution of estimates was provided, we derived the standard deviation from the parametric distribution. Otherwise, we used the 95%CI, if available, to derive the standard deviation using the formula 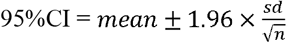.

For studies reporting only the median and its associated dispersion measure, the corresponding mean, standard deviation, and 95%CI were approximated using methods described in Luo et al. [27]. If a study did not include a measure of central tendency or dispersion, the study was included in the review, but not in the meta-analysis. These methods were implemented using R function *metagen* from package *meta*. The pooled average estimates with 95%CI were shown in forest plots.

We used a weighted t-test to assess the significance of the difference between the incubation periods, serial intervals, and generation times of the ancestral lineage and the Alpha, Delta, and Omicron variants. Weights were estimated as the inverse of the standard error, and a 2-sided p-value was calculated from a t-distribution using a bootstrap procedure to account for non-normality [28] [29].

Publication bias was assessed using a funnel plot and Egger’s test. A 2-sided P <0.05 was considered statistically significant. All analyses were performed in R (version 4.1.0.). This review was not registered.

## Results

### Search results

A total of 25,929 studies were identified based on our search strategy. After excluding 3,259 duplicated studies and an additional 22,424 articles via screening titles and abstracts, 246 articles were assessed for eligibility through a meticulous review of the full article. As per the inclusion/exclusion criteria, 147 studies were included in our analysis [7, 8, 17, 26, 30-172], 7 of which were from preprint platforms. Among the included studies, 92 studies provided incubation period estimates, 98 studies provided serial interval estimates, and 21 studies provided generation time estimates. Sixty-three studies reported more than one outcome; 18 studies provided estimates for more than one virus lineage (Figure 1).

**Figure 1.**
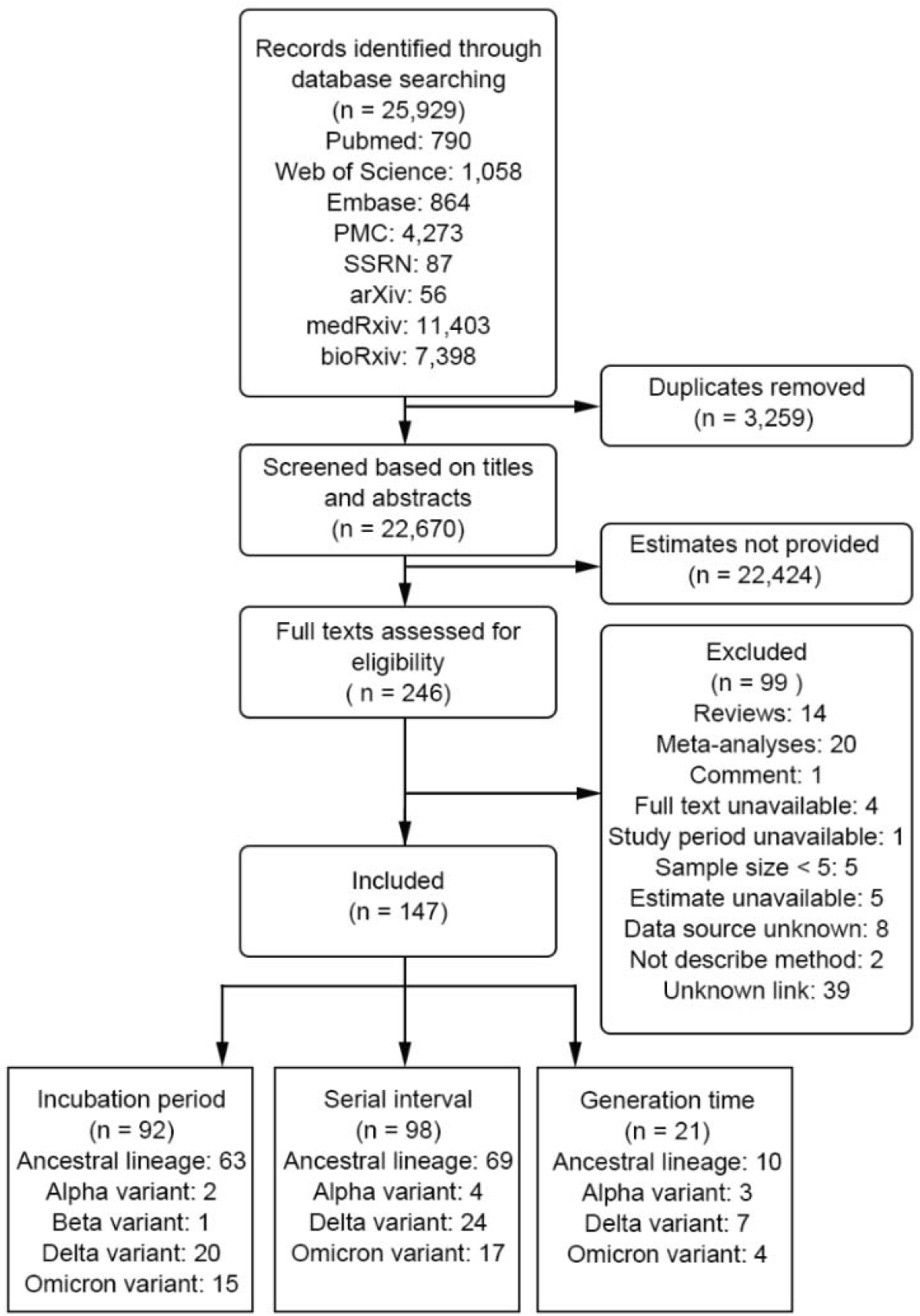
Study Flow Diagram.

### Study characteristics

We extracted 274 records for the focus key time-to-event periods from 147 included studies. Each record contained at least one summary statistic of central tendency and dispersion measure of the outcome variable(s) (Table S4-S7 in the Supplement).

We identified 109 (39.8%) records for incubation period from 92 studies; the majority of which were based on COVID-19 data from China (66 [60.6%]). Sixty-nine (63.3%) studies focused on the ancestral lineage, 2 (1.8%) on the Alpha variant, 1 (0.9%) on the Beta variant, 20 (18.3%) on the Delta variant, and 17 (15.6%) on the Omicron variant.

For serial interval, we obtained 133 (48.5%) records from 99 studies. Nearly half of the records were from China (46 [34.6%]) and South Korea (14 [10.5%]). Seventy-one (53.4%) studies included focused on the ancestral lineage, 4 (3.0%) on the Alpha variant, 29 (21.8%) on the Delta variant, and 29 (21.8%) on the Omicron variant.

For the generation time, 32 (11.7%) records from 21 studies were included in our analysis, among them were 27 records for the realized generation time and 5 records for intrinsic generation time. These records were from China (12 [37.5%]), Italy (7 [21.9%]), UK (5 [15.6%]), Netherlands (4 [12.5%]), Singapore (2 [6.3%]), Germany (1 [3.1%]), and multiple countries (1 [3.1%]). Eleven (34.4%) studies focused on the ancestral lineage, 5 (15.6%) on the Alpha variant, 10 (31.3%) on the Delta variant, and 6 (18.8%) on the Omicron variant.

Quality assessment (Table S3 in the Supplement) indicated that 1 study provided a precise exposure window for cases and identification of the potential infector(s). Sixty-eight studies included a well-characterized cohort of individuals that were comparable with the population and provided precise estimates for the symptom onset window for themselves and their potential infector(s).

### Incubation period

The ancestral lineage had the largest pooled mean incubation period (6.46 days, 95%CI: 5.86-7.07 days), followed by the Alpha variant (4.92 days, 95%CI: 4.53-5.30 days), the Delta variant (4.63, 95%CI: 4.11-5.15 days), and the Omicron variant (3.63 days, 95%CI: 3.25-4.02 days) (Figure 2, Figure S1). Only one study reported an estimate for the incubation period of the Beta variant (median: 4.5 days, IQR: 2-7 days). Each VOC had a significantly shorter incubation period than the ancestral lineage (p-values: <0.001, <0.001, <0.001 for Alpha, Delta, and Omicron, respectively). Over the course of the pandemic, the central tendency of the incubation period decreased by 23.8% from the ancestral lineage to Alpha (p-value: <0.001), 21.5% from Delta to Omicron (p-value: 0.006), and there was no significant difference between Alpha and Delta (p-value: 0.321). Our results suggested no potential publication bias in the included studies (p-value: 0.0769) (Figure S5 in the Supplement).

**Figure 2.**
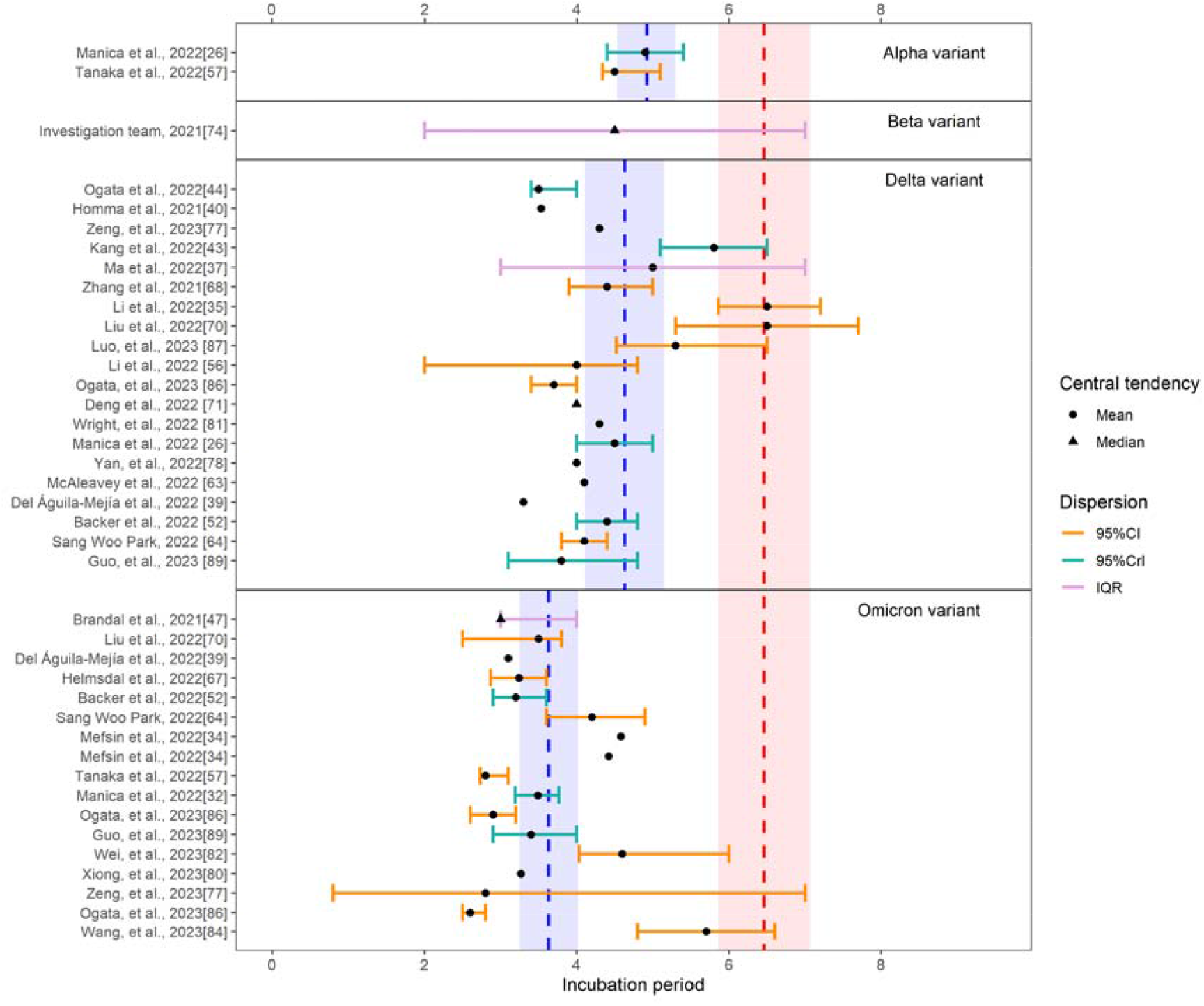
The reported estimates of the incubation period. The red vertical dotted line and rectangle correspond to the pooled mean estimate and its 95%CI of the ancestral lineage, respectively. The blue vertical dotted line and rectangle in different strata denote the pooled mean estimates and their 95%CI of corresponding variants, respectively. Black points and triangles represent mean estimates and median estimates, respectively. The horizontal segments indicate CI (orange), CrI (green), and IQR (purple). Abbreviations: CI, confidence interval; CrI, credible interval; IQR, interquartile range.

### Serial interval

The ancestral lineage had the largest pooled mean serial interval (4.80 days, 95%CI: 4.48-5.11 days), followed by the Alpha variant (3.47 days, 95%CI: 2.52-4.41 days), the Delta variant (3.59 days, 95%CI: 3.26-3.92 days) and the Omicron variant (3.19 days, 95%CI: 2.95-3.43 days) (Figure 3, Figure S2). A significantly shorter serial interval was found for each VOC compared to the ancestral lineage (p-values: 0.032, <0.001, <0.001 for Alpha, Delta, and Omicron, respectively). Likewise, the central tendency of the serial interval subsequently decreased by 27.7% from the ancestral lineage to Alpha (p-value: 0.032), 11.1% from the Delta to Omicron (p-value: 0.024). And the differences between Alpha and Delta (p-value: 0.602) was not significant. Our results suggested no potential publication bias in the included studies (p-value: 0.764) (Figure S6 in the Supplement).

**Figure 3.**
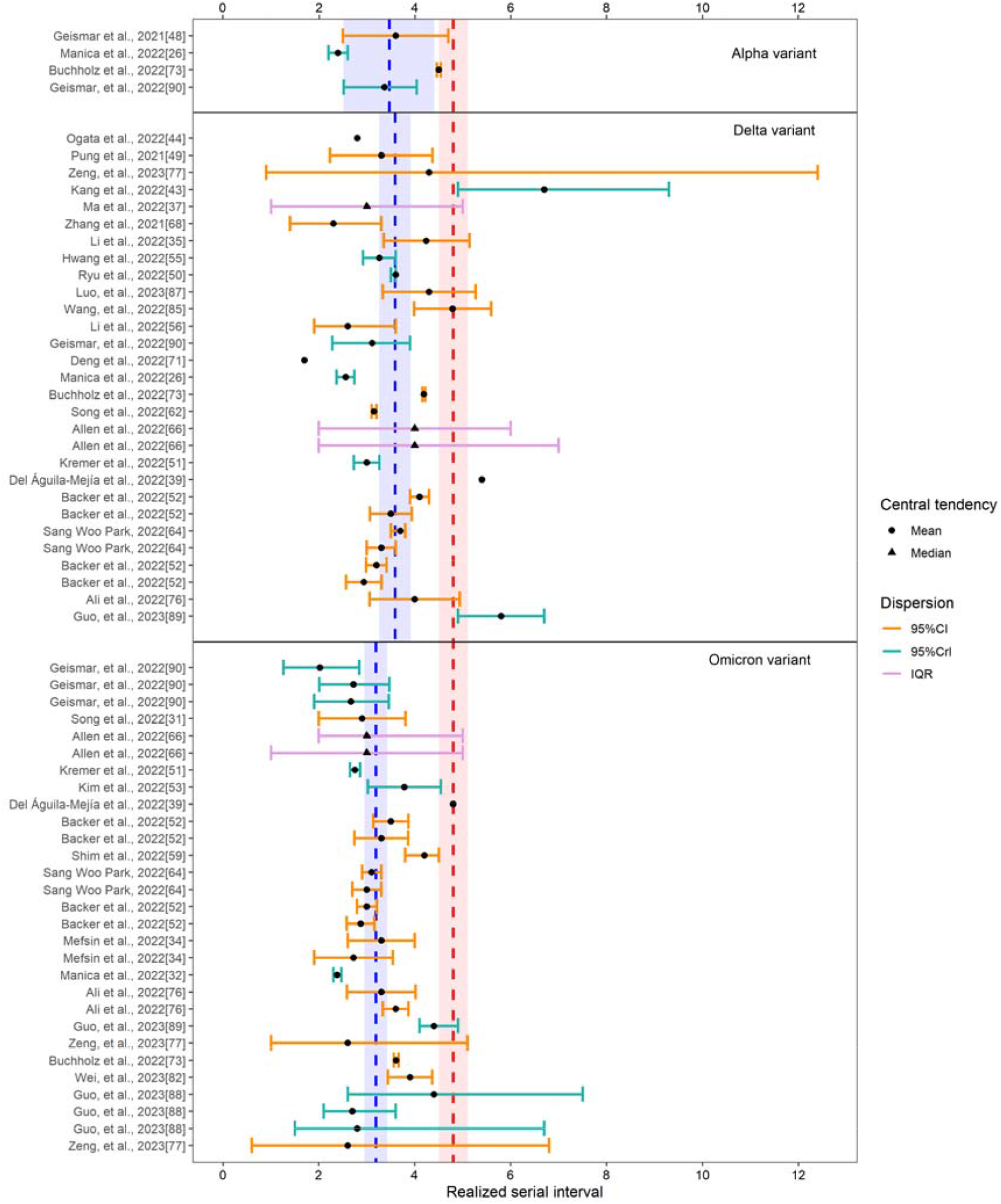
The reported estimates of the realized serial interval. The red vertical dotted line and rectangle correspond to the pooled mean estimate and its 95%CI of the ancestral lineage, respectively. The blue vertical dotted line and rectangle in different strata denote the pooled mean estimates and their 95%CI of corresponding variants, respectively. Black points and triangles represent mean estimates and median estimates, respectively. The horizontal segments indicate CI (orange), CrI (green), and IQR (purple). Abbreviations: CI, confidence interval; CrI, credible interval; IQR, interquartile range.

### Generation time

For the realized generation time, the ancestral lineage had the largest pooled mean generation time (4.94 days, 95%CI: 4.29-5.58 days), followed by the Alpha variant (4.36 days, 95%CI: 3.91-4.80 days), the Delta variant (3.65 days, 95%CI: 3.25-4.05 days), and the Omicron variant (2.96 days, 95%CI: 2.54-3.38 days) (Figure 4, Figure S3). The realized generation times for the Delta and Omicron variants were significantly shorter than the ancestral lineage (p-values: 0.005, <0.001 for Delta and Omicron, respectively). The central tendency of the generation time subsequently decreased by 16.3% from Alpha to Delta (p-value: 0.028), 18.9% from Delta to Omicron (p-value: 0.043), and there was no significant difference between ancestral lineage and Alpha (p-value: 0.193). Our results suggested no potential publication bias in the included studies (p-value: 0.709) (Figure S7 in the Supplement).

**Figure 4.**
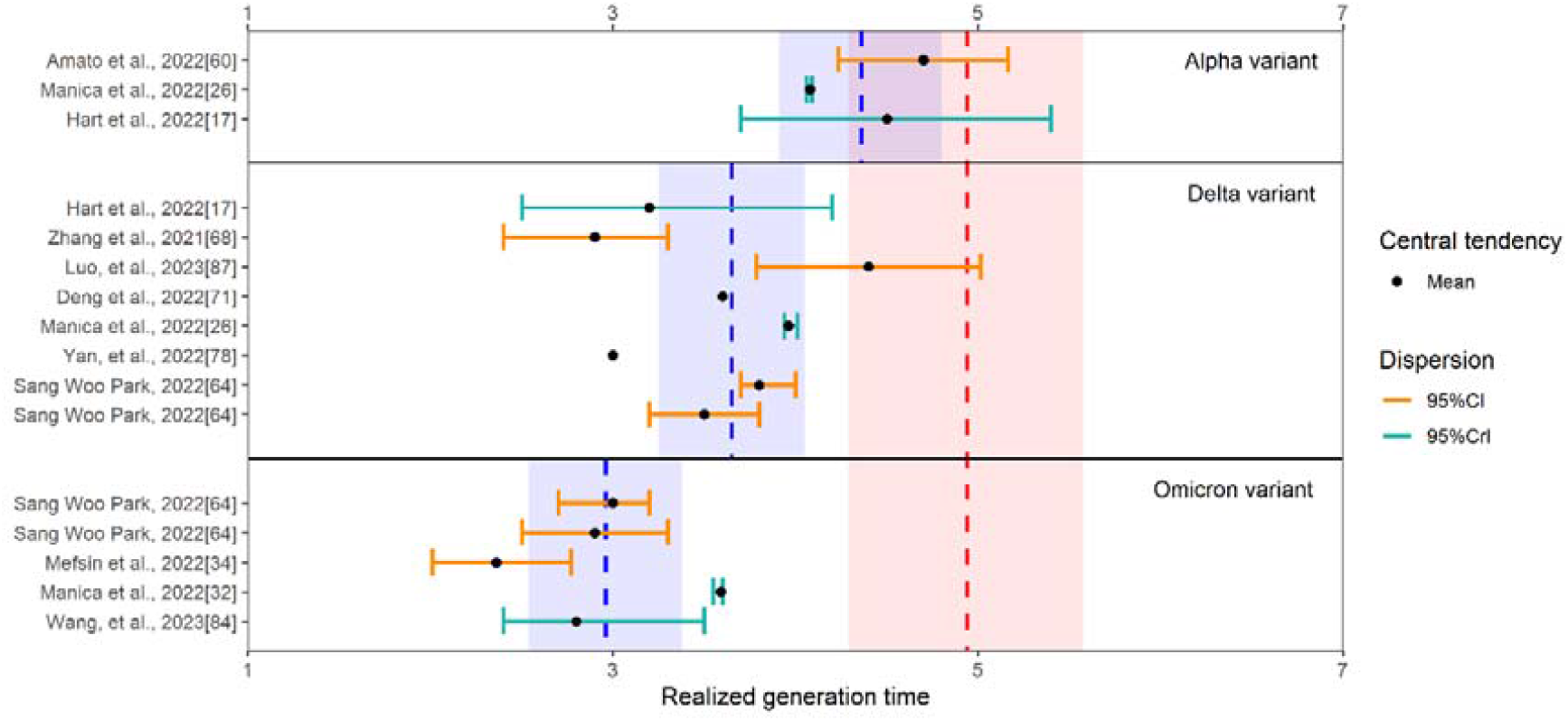
The reported estimates of the realized generation time. The red vertical dotted line and rectangle correspond to the pooled mean estimate and its 95%CI of the ancestral lineage, respectively. The blue vertical dotted line and rectangle in different strata denote the pooled mean estimates and their 95%CI of corresponding variants, respectively. Black points and triangles represent mean estimates and median estimates, respectively. The horizontal segments indicate CI (orange), CrI (green), and IQR (purple). Abbreviations: CI, confidence interval; CrI, credible interval; IQR, interquartile range.

For the intrinsic generation time, we obtained a pooled mean estimate of 5.86 days (95%CI: 5.47-6.26 days) for the Alpha variant based on two studies and 5.67 days (95%CI: 3.79-7.55 days) for the Delta variant based on two studies (Figure 5, Figure S4). Only one study (for Italy) reported an estimate for the Omicron variant: 6.84 days (95%CrI: 5.72-8.60 days).

**Figure 5.**
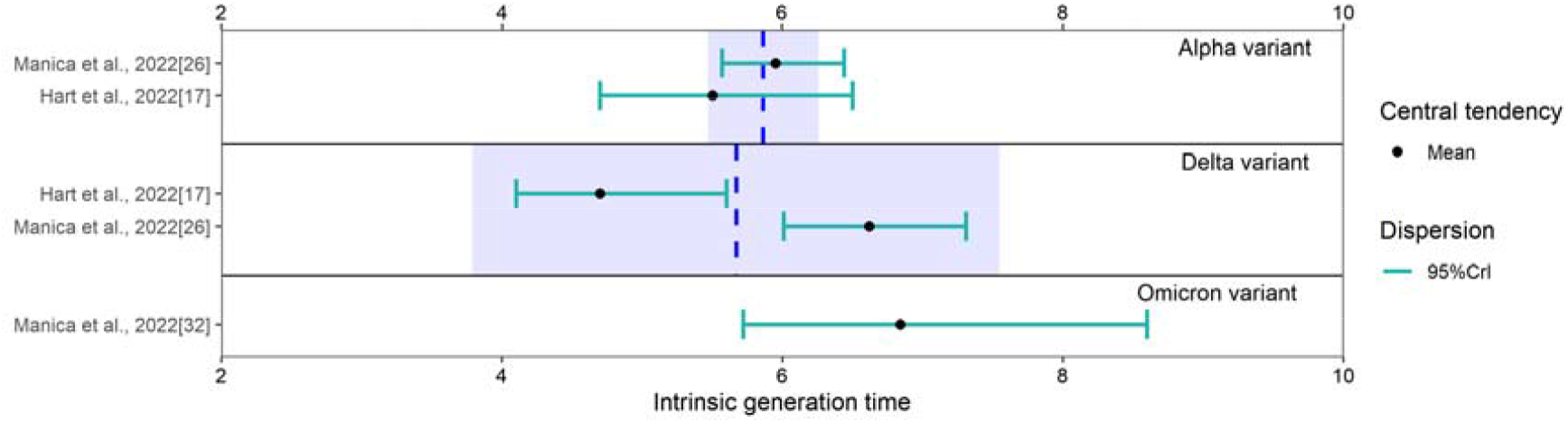
The reported estimates of the intrinsic generation time. The blue vertical dotted line and rectangle in different strata denote the pooled mean estimates and their 95%CI of corresponding variants, respectively. Black points and triangles represent mean estimates and median estimates, respectively. The horizontal segments indicate CI (orange), CrI (green), and IQR (purple). Abbreviations: CI, confidence interval; CrI, credible interval; IQR, interquartile range.

## Discussion

Our study revealed that Omicron had the shortest pooled estimates for the incubation period (3.63 days, 95%CI: 3.25-4.02 days), serial interval (3.19 days, 95%CI: 2.95-3.43 days), and realized generation time (2.96 days, 95%CI: 2.54-3.38 days) compared to previous variants. Our results were comparable to those of Galmiche, S., et al., who reported SARS-CoV-2 incubation period was notably reduced in omicron cases compared with all other variants of concern [173]. The ancestral lineage had the highest pooled estimates for each of the three indicators, followed by Alpha and Delta. These finding suggest that the incubation period, serial interval, and realized generation times of COVID-19 became shorter over the course of the pandemic.

The majority of the studies included in our analysis provided estimates for the incubation period (92 studies) and serial interval (99 studies), while only 21 studies provided estimates for the generation time. This suggests that estimates for serial interval and incubation period are easier to obtain as they can more easily inferred from contact tracing and household studies than estimates for generation time, which requires more complex Bayesian analyses as the date of infection of the infector and their infectees are both generally unknown [26, 32]. Furthermore, China provided more records of estimates for incubation period (60.6%), serial interval (34.6%), and generation time (37.5%) than any other country. This may be due to the smaller outbreak size of COVID-19 outbreaks in China before the rise of Omicron as compared to countries with widespread COVID-19 transmission, which made contact tracing and household studies easier to be conducted. This further support the importance of contact tracing not only as a tool for monitoring and controlling infectious disease spread, but also for understanding transmission patterns.

Examining the intrinsic generation time is done using recently developed Bayesian methods, which may explain why only two studies have provided estimates for this indicator [17, 32]. Between these studies, there were only 5 records: 2 for Alpha, 2 the Delta, and 1 for Omicron. Given the small sample size, a pooled mean estimate was not warranted, and we could not compare possible differences between subsequent VOCs.

The incubation period of the ancestral lineage, and the Alpha, Beta, and Delta variants, is generally longer than that of other acute respiratory viral infections, such as human coronavirus (3.2 days), influenza A (1.43-1.64 days), parainfluenza (2.6 days), respiratory syncytial virus (4.4 days), and rhinovirus (1.4 days) [174]. Our findings produced similar mean incubation period estimates to those reported by Du, Z., et al., for Delta (4.8 days 95% CI: 3.9-5.6) and Omicron (3.6 days, 95% CI: 2.3-4.9) [175] and those reported by Wu et. al., for Alpha (5.00 days, 95% CI: 4.94-5.06 days), Delta (4.41 days, 95% CI: 3.76-5.05 days), and Omicron (3.42 days, 95% CI: 2.88-3.96 days) [23].

Our study showed that the serial interval of COVID-19 ranged from 1.7 to 7.5 days. These estimates are longer than those of influenza A(H3N2) (2.2 days) and pandemic influenza A(H1N1)pdm09 (2.8 days), but shorter than those of the respiratory syncytial virus (RSV, 7.5 days), measles (11.7 days), varicella (14.0 days), smallpox (17.7 days), mumps (18.0 days), rubella (18.3 days), pertussis (22.8 days), and Middle East respiratory syndrome (MERS, 7.6 days) [176] [177]. Our results were comparable to those of Du, Z., et al., who reported an average serial interval of 3.4 days (95% CI, 3.0-3.7) for the Delta variant and 3.1 days (95% CI, 2.9-3.2) for the Omicron variant, respectively [175].

The mean generation time of COVID-19 ranged from 2.36 to 6.84 days, longer than those of influenza A (H1N1) (2 days) and pandemic influenza A(H1N1)pdm09 (2.92 days) [178], but shorter than those of the MERS (10.7 days) [179, 180].

Overall, the serial interval maintained a shorter pooled mean estimate compared to the incubation period across different virus lineages, indicating that a large proportion of SARS-CoV-2 transmission occurs prior to symptom onset [181-183]. Pre-symptomatic transmission was a critical factor facilitating SARS-CoV-2 spread [184], highlighting the importance of obtaining timely estimates of these indicators and keep monitoring their possible changes over the course of an epidemic.

The current study provides scientific evidence that the incubation period of COVID-19 has shortened as the virus has evolved, which has important implications for the formulation of effective epidemic control strategies such as isolation and quarantine. It is still unclear whether the increase of immunity levels in the population has contributed to this trend, as prior infection or vaccination can affect the incubation period by altering the viral load and shedding [185]. Similarly, it is unclear whether the diverse immunological landscape in different countries has contributed to the substantial heterogeneity of the incubation period observed for the same variant across different studies.

Our findings also demonstrated that the serial intervals of COVID-19 shortened with each new VOC. Previous studies have attributed decreased serial intervals to preventive measures that target the duration of potential transmission, such as isolation, contact tracing, quarantine, and other non-pharmaceutical interventions (NPIs) [10-12, 49, 186], which is further supported by the fact that longer serial intervals are often censored due to case isolations [35]. The highly heterogeneous implementation of NPIs between countries, within country, and time frame has probably contributed to heterogeneous estimates of the serial interval that we observed between studies. This applies also for estimates of the realized generation time.

We acknowledge some limitations in this study. First, results of the included studies revealed considerable heterogeneities (I^2^ > 80%) when pooling the estimates across different virus lineages, indicating potential unmeasured confounding from population factors (e.g., social behavior). Specifically, heterogeneities for the incubation period may be due to heterogeneities in the age structure and presence of pre-existing conditions in the host population [187, 188], whereas diversity in contact settings and the strength of NPIs may be responsible for heterogeneity of the estimates for the serial interval and realized generation time. However, these data were rarely available from the included published literature and preprints, thus preventing further exploration in our study. As per the inclusion criteria, we only used data collected from studies published in English potentially subjecting our study to reporting bias, which may impact our results. This study may be limited by recall bias as many studies included in the analysis rely on retrospective data collection for exposure and symptom onset, which would influence the obtained estimates. The incubation period for the Beta variant and the intrinsic generation time for Omicron were each included in only one study, and thus it was not possible to produce pooled estimates.

In conclusion, our findings suggest that the incubation period, serial interval, and generation time of SARS-CoV-2 have evolved to shorter intervals with the emergence of each new VOC. Identifying the length of each of these indicators is critical for understanding the epidemiology of different SARS-CoV-2 variants and developing control measures for mitigating the spread of COVID-19. Moreover, understanding trends in these indicators can be instrumental for preparedness planning for future COVID-19 outbreaks.

## Data Availability

All data are collected from open source with detailed description in Supplemental Information.

## Declarations

### Authors’ contributions

M.A., YP.W., Y.W., H.L. conceived and designed the study. H.Y., and M.A. supervised the study. X.X., YP.W., Y.Z. and Z.H. participated in data collection. X.X. and YP.W. participated in statistical analysis. X.X. and Y.Z. prepared the tables and figures. YP.W., A.G.K. and X.X. drafted the manuscript. M.A., A.G.K. and YP.W. revised the content critically. All authors contributed to review and revision and approved the final manuscript as submitted and agree to be accountable for all aspects of the work.

### Availability of data

All data are collected from open source with detailed description in Supplemental Information.

### Ethics approval and consent to participate

Not applicable.

## Acknowledgements

Not applicable.

## Consent for publication

Not applicable.

