## Supplemental Table S1-S8, Figure S1-S7 for "Assessing changes in incubation period, serial interval, and generation time of SARS-CoV-2 variants of concern: a systematic review and meta-analysis"

*Joint senior authors

### Table S1. The details of the article search strategies and search results for epidemiological parameters using search terms specific to the corresponding database.

| **Database** | **Step** | **Search strategy** | **Number of articles*** |
| --- | --- | --- | --- |
| PubMed | #1 | COVID-19 OR SARS-CoV-2 OR 2019-nCoV OR "coronavirus disease 2019" OR "severe acute respiratory syndrome coronavirus 2" | 348,819 |
|  | #2 | ("serial interval*" OR "generation time*" OR "generation interval*") AND (contact tracing OR household) | 196 |
|  | #3 | "incubation period*" | 15,603 |
|  | #4 | English [Language] | 30,661,800 |
|  | #5 | #1 AND #2 AND #4 | 108 |
|  | #6 | #1 AND #3 AND #4 | 725 |
|  | #7 | #5 OR #6 | 790 |
| Embase | #1 | (COVID-19 OR SARS-CoV-2 OR 2019-nCoV OR "coronavirus disease 2019" OR "severe acute respiratory syndrome coronavirus 2") AND ((("serial interval*" OR "generation time*" OR "generation interval*") AND (contact tracing OR household)) OR "incubation period*") | 864 |
|  | #1 | TS = (COVID-19 OR SARS-CoV-2 OR 2019-nCoV OR "coronavirus disease 2019" OR "severe acute respiratory syndrome coronavirus 2") | 520,222 |
| Web of Science | #2 | TS = (("serial interval*" OR "generation time*" OR "generation interval*") AND (contact tracing OR household)) | 263 |
|  | #3 | TS = ("incubation period*") | 40,802 |
|  | #4 | #1 AND #2 | 137 |
|  | #5 | #1 AND #3 | 1,167 |
|  | #6 | #4 OR #5 | 1,244 |
|  | #7 | #6 AND Language:English | 1,058 |
| Europe PMC | #1 | (COVID-19 OR SARS-CoV-2 OR 2019-nCoV OR "coronavirus disease 2019" OR "severe acute respiratory syndrome coronavirus 2") AND ((("serial interval" OR "generation time" OR "generation interval” OR "serial intervals" OR "generation times" OR "generation intervals") AND (contact tracing OR household)) OR “incubation periods") AND (LANG:"eng" OR LANG:"en" OR LANG:"us") | 4,273 |
| medRxiv | #1 | SARS-CoV-2 AND "serial interval*" AND contact tracing | 250 |
|  | #2 | SARS-CoV-2 AND "generation time*" AND contact tracing | 8,502 |
|  | #3 | SARS-CoV-2 AND "generation interval*" AND contact tracing | 862 |
|  | #4 | SARS-CoV-2 AND "incubation period*" | 4,871 |
|  | #5 | SARS-CoV-2 AND "serial interval*" AND household | 150 |
|  | #6 | SARS-CoV-2 AND "generation time*" AND household | 3,691 |
|  | #7 | SARS-CoV-2 AND "generation interval*" AND household | 452 |
|  | #8 | #1 OR #2 OR #3 OR #4 OR #5 OR #6 OR #7 | 11,403 |
| bioRxiv | #1 | SARS-CoV-2 AND "serial interval*" AND contact tracing | 31 |
|  | #2 | SARS-CoV-2 AND "generation time*" AND contact tracing | 3,515 |
|  | #3 | SARS-CoV-2 AND "generation interval*" AND contact tracing | 77 |
|  | #4 | SARS-CoV-2 AND "incubation period*" | 5,940 |
|  | #5 | SARS-CoV-2 AND "serial interval*"AND household | 6 |
|  | #6 | SARS-CoV-2 AND "generation time*" AND household | 298 |
|  | #7 | SARS-CoV-2 AND "generation interval*" AND household | 17 |
|  | #8 | #1 OR #2 OR #3 OR #4 OR #5 OR #6 OR #7 | 7,398 |
| arXiv | #1 | COVID-19 OR SARS-CoV-2 OR 2019-nCoV OR "coronavirus disease 2019" OR "severe acute respiratory syndrome coronavirus 2" | 7,123 |
|  | #2 | "serial interval" OR "generation time" OR "generation interval" OR "incubation period" OR "serial intervals" OR "generation times" OR "generation intervals" OR "incubation periods" | 1,169 |
|  | #3 | #1 AND #2 | 56 |
| SSRN | #1 | COVID-19 AND "serial interval*" | 14 |
|  | #2 | COVID-19 AND "generation time*" | 6 |
|  | #3 | COVID-19 AND "generation interval*" | 1 |
|  | #4 | COVID-19 AND "incubation period*" | 39 |
|  | #5 | SARS-CoV-2 AND "serial interval*" | 8 |
|  | #6 | SARS-CoV-2 AND "generation time*" | 5 |
|  | #7 | SARS-CoV-2 AND "generation interval*" | 1 |
|  | #8 | SARS-CoV-2 AND "incubation period*" | 13 |

*Searches were conducted on 28 March 2023.

### Table S2. Quality assessment scale

| A | **Robustness of data collection for contacts**   1. Includes contacts regardless of the clinical outcome, including individuals that test negative 2. Includes contacts from a specific setting (e.g., household) regardless of the clinical outcome, including individuals that test negative 3. Includes a subset of contacts that tested positive 4. No description/not clear |
| --- | --- |
| B | **Representativeness of the study cohort – Applies to all estimates**   1. No selection of cases based on age, sex, or general health status, supported by descriptive statistics demonstrating comparability with overall population 2. No selection of cases based on age, sex, or general health status, not supported by descriptive statistics 3. Cases are likely to be biased towards those with more severe COVID-19 symptoms due to selection process – e.g. records from hospitalized patients 4. Cases are selected (e.g. based on age or sex) to represent a particular cohort of individuals 5. No description of the derivation of the cohort |
| C | **Precision of the exposure window for cases used in final analysis – Applies to incubation period and generation time estimates**   1. Only includes cases with a 1-day exposure window 2. Only includes cases with less than or equal to 3-day exposure window 3. Includes cases with more than 3-day exposure window/unknown length of window 4. Includes cases with first day, last day or intermediate day of exposure window 5. No description/not clear |
| D | **Identification of potential infector(s) – Applies to all estimates**   1. Multiple exposures but statistical methods are applied to account for this 2. Multiple exposures but no statistical methods are applied to account for this 3. Single exposure 4. No description/not clear |
| E | **Precision of estimate of the symptom onset – Applies to incubation period and serial interval estimates**   1. Precise date 2. Window 3. No description/not clear |
| F | **Precision of estimate for symptom onset of identified potential infector(s) – Applies to serial interval estimate**   1. Precise date 2. Window 3. No description/not clear |
| G | **Distribution of the incubation period used in the analysis – Applies to generation time estimate**   1. Both the variant and location match those used in the estimation. 2. Either the variant or the location match those used in the estimation. 3. Neither the variant nor the location match those used in the estimation. 4. No description/not used |

### Table S3. Quality assessment of final studies used in the meta-analysis of incubation period, serial interval, and generation time

|  | Study | Quality Assessment | | | | | | |
| --- | --- | --- | --- | --- | --- | --- | --- | --- |
|  |  | A | B | C | D | E | F | G |
| Ancestral Lineage | Shen et al., 2020[1] | 1 | 1 | 4 | 4 | 1 | NA | NA |
|  | Vazirinejad et al., 2020[2] | 1 | 1 | NA | 4 | 1 | 1 | NA |
|  | Bender et al., 2021[3] | 1 | 1 | 3 | 4 | 1 | 1 | 4 |
|  | Liu et al., 2020[4] | 3 | 1 | 5 | 4 | 1 | 1 | NA |
|  | Haddad et al., 2021[5] | 1 | 1 | NA | 4 | 1 | 1 | NA |
|  | Shi et al., 2020[6] | 1 | 1 | 5 | 4 | 1 | 1 | NA |
|  | Song et al., 2020[7] | 2 | 1 | 5 | 4 | 3 | NA | NA |
|  | Gupta et al., 2020[8] | 1 | 1 | NA | 4 | 1 | 1 | NA |
|  | Bao et al., 2021[9] | 1 | 1 | 5 | 4 | 1 | 1 | NA |
|  | Du et al., 2020[10] | 3 | 1 | NA | 4 | 1 | 1 | NA |
|  | Li et al., 2020[11] | 3 | 1 | 5 | 4 | 1 | 1 | NA |
|  | Zhu et al., 2021[12] | 3 | 1 | 3 | 4 | 1 | 1 | NA |
|  | Ping et al., 2021[13] | 1 | 1 | 3 | 4 | 1 | 1 | NA |
|  | Ki,2020[14] | 1 | 1 | 5 | 4 | 1 | 1 | NA |
|  | Mao et al., 2020[15] | 1 | 1 | 1 | 3 | 1 | NA | NA |
|  | Zhang et al., 2020[16] | 1 | 1 | 5 | 4 | 1 | NA | NA |
|  | Guo et al., 2020[17] | 3 | 4 | 5 | 4 | 1 | NA | NA |
|  | Nie et al., 2020[18] | 3 | 1 | 4 | 4 | 1 | NA | NA |
|  | Son et al., 2020[19] | 1 | 1 | NA | 4 | 1 | 1 | NA |
|  | Baskaradoss et al., 2021[20] | 1 | 1 | NA | 4 | 1 | 1 | NA |
|  | Du et al., 2021[21] | 3 | 1 | 4 | 4 | 1 | NA | NA |
|  | Thway et al., 2020[22] | 3 | 1 | NA | 4 | 1 | 1 | NA |
|  | Kwok et al., 2020[23] | 1 | 1 | NA | 4 | 1 | 1 | NA |
|  | Hua et al., 2020[24] | 2 | 4 | 4 | 4 | 1 | NA | NA |
|  | Wong et al., 2020[25] | 1 | 1 | 1 | 4 | 1 | 1 | NA |
|  | Haw et al., 2020[26] | 3 | 1 | NA | 4 | 1 | 1 | NA |
|  | Ganyani et al., 2020[27] | 3 | 2 | 5 | 4 | NA | NA | 2 |
|  | Hart et al., 2022[28] | 2 | 1 | 5 | 1 | NA | NA | 1 |
|  | Hu et al., 2021[29] | 1 | 1 | 4 | 4 | NA | NA | 2 |
|  | Zhao et al., 2021[30] | 3 | 2 | 5 | 4 | 1 | NA | 4 |
|  | Lau et al., 2021[31] | 3 | 2 | 3 | 4 | 1 | NA | 4 |
|  | Deng et al., 2021[32] | 3 | 2 | 3 | 4 | 1 | NA | NA |
|  | Böhm et al., 2021[33] | 1 | 1 | 3 | 3 | 1 | 1 | NA |
|  | Böhmer et al., 2020[34] | 1 | 1 | 4 | 4 | 1 | 1 | NA |
|  | Yang et al., 2020[35] | 3 | 1 | 4 | 3 | 1 | 1 | NA |
|  | Zhang et al., 2020[36] | 3 | 1 | 3 | 3 | 1 | 1 | NA |
|  | Backer et al., 2020[37] | 4 | 1 | 3 | 4 | 1 | NA | NA |
|  | Bui et al., 2020[38] | 3 | 1 | 3 | 4 | 1 | NA | NA |
|  | Cheng et al., 2021[39] | 4 | 1 | 3 | 4 | 1 | NA | NA |
|  | Han,2020[40] | 1 | 1 | 5 | 4 | 1 | NA | NA |
|  | Kong,2020[41] | 4 | 2 | 2 | 4 | 1 | NA | NA |
|  | Xiao et al., 2021[42] | 4 | 1 | 5 | 4 | 1 | NA | NA |
|  | Xiao et al., 2020[43] | 4 | 1 | 5 | 4 | 1 | NA | NA |
|  | Ratovoson et al., 2022[44] | 2 | 1 | 3 | 4 | 1 | 1 | NA |
|  | Aghaali et al., 2020[45] | 1 | 2 | NA | 4 | 3 | 3 | NA |
|  | Bi et al., 2020[46] | 1 | 1 | 3 | 4 | 1 | 1 | NA |
|  | Expert Taskforce,2020[47] | 1 | 1 | NA | 4 | 1 | 1 | NA |
|  | McAloon et al., 2021[48] | 1 | 2 | NA | 4 | 3 | 3 | NA |
|  | Talmoudi et al., 2020[49] | 3 | 2 | NA | 4 | 1 | 1 | NA |
|  | Wang et al., 2020[50] | 3 | 1 | NA | 4 | 1 | 1 | NA |
|  | Kwok et al., 2021[51] | 3 | 1 | NA | 3 | 1 | 1 | NA |
|  | Dai et al., 2020[52] | 3 | 1 | 3 | 4 | 1 | NA | NA |
|  | Huang et al., 2020[53] | 1 | 4 | 5 | 4 | 1 | 1 | NA |
|  | Areekal et al., 2021[54] | 1 | 1 | 4 | 4 | 1 | 1 | NA |
|  | Tindale et al., 2020[55] | 3 | 2 | 3 | 4 | 1 | 1 | NA |
|  | Won et al., 2021[56] | 3 | 2 | 5 | 4 | 1 | 1 | NA |
|  | Xia et al., 2020[57] | 3 | 1 | 2 | 4 | 1 | 1 | NA |
|  | Zhao et al., 2021[58] | 1 | 1 | 4 | 4 | 1 | 1 | NA |
|  | Emecen et al., 2021[59] | 3 | 3 | 3 | 3 | 1 | 1 | NA |
|  | Li et al., 2020[60] | 3 | 2 | 4 | 4 | 1 | 1 | 4 |
|  | Ali et al., 2020[61] | 3 | 2 | NA | 4 | 1 | 1 | NA |
|  | Bernal Lopez et al., 2022[62] | 2 | 1 | 5 | 4 | 1 | 1 | NA |
|  | Cereda et al., 2021[63] | 1 | 1 | NA | 4 | 1 | 1 | NA |
|  | Hong et al., 2020[64] | 3 | 1 | NA | 4 | 3 | 3 | NA |
|  | Lavezzo et al., 2020[65] | 2 | 1 | NA | 4 | 3 | 3 | NA |
|  | Liu et al., 2020[66] | 3 | 2 | NA | 4 | 1 | 1 | NA |
|  | Najafi et al., 2020[67] | 3 | 2 | NA | 4 | 1 | 1 | NA |
|  | Prete et al., 2020[68] | 3 | 2 | NA | 4 | 1 | 1 | NA |
|  | Reed et al., 2021[69] | 3 | 2 | NA | 4 | 1 | 1 | NA |
|  | Ryu et al., 2021[70] | 3 | 2 | NA | 4 | 1 | 1 | NA |
|  | Saurabh et al., 2020[71] | 3 | 2 | NA | 4 | 1 | 1 | NA |
|  | Thai et al., 2021[72] | 1 | 2 | NA | 4 | 1 | 1 | NA |
|  | Qin et al., 2020[73] | 4 | 1 | 5 | 4 | 1 | NA | NA |
|  | Ren et al., 2021[74] | 3 | 1 | 3 | 4 | 1 | 1 | NA |
|  | Zhao et al., 2020[75] | 3 | 2 | NA | 4 | 1 | 1 | NA |
|  | Adam et al., 2020[76] | 3 | 2 | NA | 3 | 1 | 1 | NA |
|  | Ferretti et al., 2020[77] | 3 | 2 | 5 | 4 | NA | NA | 2 |
|  | Althobaity et al., 2022[78] | 3 | 2 | 3 | 4 | 1 | 1 | NA |
|  | Wang et al., 2022[79] | 1 | 2 | NA | 4 | 1 | 1 | NA |
|  | Geismar et al., 2021[80] | 2 | 1 | NA | 3 | 1 | 1 | NA |
|  | Buchholz et al., 2022[81] | 2 | 2 | NA | 3 | 1 | 1 | NA |
|  | Somda, et al., 2022 [82] | 3 | 1 | NA | 4 | 3 | 3 | NA |
|  | Hammond, et al., 2022 [83] | 1 | 1 | 3 | 3 | 1 | 1 | NA |
|  | Geismar, et al., 2022 [84] | 2 | 2 | NA | 4 | 3 | 3 | NA |
| Alpha | Hart et al., 2022[85] | 2 | 2 | 5 | 1 | NA | NA | 3 |
|  | Manica et al., 2022[86] | 2 | 1 | 3 | 1 | 1 | 1 | 1 |
|  | Geismar et al., 2021[80] | 2 | 1 | NA | 3 | 1 | 1 | NA |
|  | Tanaka et al., 2022[87] | 1 | 1 | 1 | 3 | 1 | NA | NA |
|  | Amato et al., 2022[88] | 3 | 1 | 3 | 2 | NA | NA | 3 |
|  | Buchholz et al., 2022[81] | 2 | 2 | NA | 3 | 1 | 1 | NA |
|  | Geismar, et al., 2022 [84] | 2 | 2 | NA | 4 | 3 | 3 | NA |
| Beta | Investigation team, 2021[89] | 1 | 1 | 5 | 4 | 1 | NA | NA |
| Delta | Hart et al., 2022[85] | 2 | 2 | 5 | 1 | NA | NA | 3 |
|  | Manica et al., 2022[86] | 2 | 1 | 3 | 1 | 1 | 1 | 1 |
|  | Li et al., 2022[90] | 3 | 1 | 1 | 4 | 1 | 1 | NA |
|  | Kang et al., 2022[91] | 1 | 1 | 3 | 4 | 1 | 1 | NA |
|  | Ogata et al., 2022[92] | 1 | 1 | 1 | 3 | 1 | 1 | NA |
|  | Pung et al., 2021[93] | 2 | 2 | NA | 4 | 3 | 3 | NA |
|  | Ryu et al., 2022[94] | 3 | 2 | NA | 2 | 1 | 1 | NA |
|  | Kremer et al., 2022[95] | 3 | 2 | NA | 4 | 1 | 1 | NA |
|  | Backer et al., 2022[96] | 3 | 2 | 3 | 3 | 1 | 1 | NA |
|  | Hwang et al., 2022[97] | 3 | 1 | NA | 4 | 1 | 1 | NA |
|  | Li et al., 2022[98] | 1 | 1 | 5 | 4 | 1 | 1 | NA |
|  | Song et al., 2022[99] | 3 | 2 | NA | 4 | 1 | 1 | NA |
|  | McAleavey et al., 2022[100] | 3 | 1 | 5 | 4 | 1 | NA | NA |
|  | Sang Woo Park, 2022[101] | 3 | 2 | 3 | 3 | 1 | 1 | NA |
|  | Zhang et al., 2021[102] | 3 | 2 | 1 | 4 | 3 | 3 | 1 |
|  | Liu et al., 2022[103] | 4 | 2 | 3 | 4 | 1 | NA | NA |
|  | Buchholz et al., 2022[81] | 2 | 2 | NA | 3 | 1 | 1 | NA |
|  | Ali et al., 2022[104] | 3 | 2 | NA | 3 | 1 | 1 | NA |
|  | Zeng, et al., 2023 [105] | 3 | 1 | 1 | 3 | 1 | 1 | NA |
|  | Wang, et al., 2022 [106] | 3 | 1 | NA | 3 | 1 | 1 | NA |
|  | Ogata, et al., 2023 [107] | 3 | 1 | 1 | 4 | 1 | NA | NA |
|  | Luo, et al., 2023 [108] | 1 | 1 | 3 | 4 | 1 | 1 | 4 |
|  | Guo, et al., 2023 [109] | 3 | 3 | 5 | 4 | 1 | 1 | NA |
|  | Geismar, et al., 2022 [84] | 2 | 2 | NA | 4 | 3 | 3 | NA |
| Omicron | Song et al., 2022[110] | 2 | 1 | 3 | 4 | 1 | 1 | NA |
|  | Manica et al., 2022[111] | 2 | 1 | 1 | 1 | 1 | 1 | 2 |
|  | Mefsin et al., 2022[112] | 3 | 2 | 1 | 4 | 1 | 1 | 4 |
|  | Brandal et al., 2021[113] | 1 | 2 | 1 | 4 | 1 | NA | NA |
|  | Kremer et al., 2022[95] | 3 | 2 | NA | 4 | 1 | 1 | NA |
|  | Backer et al., 2022[96] | 3 | 2 | 3 | 3 | 1 | 1 | NA |
|  | Kim et al., 2022[114] | 1 | 1 | NA | 4 | 1 | 1 | NA |
|  | Tanaka et al., 2022[87] | 1 | 1 | 1 | 3 | 1 | NA | NA |
|  | Shim et al., 2022[115] | 3 | 1 | NA | 4 | 1 | 1 | NA |
|  | Sang Woo Park, 2022[101] | 3 | 2 | 3 | 3 | 1 | 1 | NA |
|  | HelmSDal et al., 2022[116] | 1 | 2 | 5 | 4 | 1 | NA | NA |
|  | Liu et al., 2022[103] | 4 | 2 | 3 | 4 | 1 | NA | NA |
|  | Buchholz et al., 2022[81] | 2 | 2 | NA | 3 | 1 | 1 | NA |
|  | Ali et al., 2022[104] | 3 | 2 | NA | 3 | 1 | 1 | NA |
|  | Zeng, et al., 2023 [105] | 3 | 1 | 1 | 3 | 1 | 1 | NA |
|  | Xiong, et al., 2023 [117] | 3 | 1 | 3 | 4 | 1 | NA | NA |
|  | Wei, et al., 2023 [118] | 2 | 3 | 3 | 3 | 3 | 3 | NA |
|  | Wang, et al., 2023 [119] | 1 | 1 | 3 | 3 | 1 | NA | 4 |
|  | Ogata, et al., 2023 [107] | 3 | 1 | 1 | 4 | 1 | NA | NA |
|  | Guo, et al., 2023 [120] | 3 | 1 | NA | 3 | 1 | 1 | NA |
|  | Guo, et al., 2023 [109] | 3 | 3 | 5 | 4 | 1 | 1 | NA |
|  | Geismar, et al., 2022 [84] | 2 | 2 | NA | 4 | 3 | 3 | NA |

Note: If a question does not apply (e.g., the study does not include generation time estimates), we classified it as NA.

### Table S4. Characteristic of included articles for incubation period and parameters extracted.

| Author | Study period | Location | Strain type | Data extracted | Data for analysis* | |
| --- | --- | --- | --- | --- | --- | --- |
|  |  |  |  |  | Mean | 95%CI |
| Shen et al., 2020 [1] | 2020.01-2020.02 | China | Ancestral lineage | n: 8 Median: 7 Range: 4-12 | 7.46 | 5.51-9.40 |
| Bender et al., 2021 [3] | 2020.02-2020.03 | Germany | Ancestral lineage | n: 53 Median: 4.3 IQR: 2.5-6.5 | 4.44 | 3.62-5.26 |
| Liu et al., 2020[4] | 2020.01-2020.04 | China | Ancestral lineage | n: 27 Mean: 6 SD: 3.1 | 6.00 | 4.83-7.17 |
| Shi et al., 2020[6] | 2020.01-2020.03 | China | Ancestral lineage | Mean: 4.77 95%CI: 3.61-5.94 | 4.77 | 3.60-5.93 |
| Zhang et al., 2021[121] | 2020.01-2020.02 | China | Ancestral lineage | Mean: 7.83 Median: 7 | / | / |
| Song et al., 2020[7] | 2020.01-2020.01 | China | Ancestral lineage | n: 22 Mean: 8.23 SD: 3.58 | 8.23 | 6.73-9.73 |
| Bao et al., 2021[9] | 2020.01-2020.02 | China | Ancestral lineage | Median: 5.4 95%CI: 4.5-6.3 | / | / |
| Li et al., 2020[11] | 2019.12-2020.01 | China | Ancestral lineage | Mean: 5.2 95%CI: 4.1-7 | 5.20 | 3.75-6.65 |
| Zhu et al., 2021[12] | 2021.01-2021.02 | China | Ancestral lineage | Mean: 11.6 95%CI: 10.6-12.7 | 11.60 | 10.55-12.65 |
| Ping et al., 2021[13] | 2020.01-2020.03 | China | Ancestral lineage | Median: 6.047 95%CI: 5.000-7.095 | / | / |
| Ki,2020[14] | 2020.01-2020.01 | South Korea | Ancestral lineage | n: 10 Median: 3 Range: 0-15 | 4.87 | 1.85-7.89 |
| Mao et al., 2020[15] | 2020.01-2020.03 | China | Ancestral lineage | n: 28 Median: 8.5 Range: 1-24 | 9.49 | 7.37-11.60 |
| Zhang et al., 2020[16] | 2020.01-2020.03 | China | Ancestral lineage | n: 8 Median: 7 IQR: 2-12 | 7.00 | 0.94-13.06 |
| Zhang et al., 2020[16] | 2020.01-2020.03 | China | Ancestral lineage | n: 23 Median: 8 IQR: 4-13 | 8.36 | 5.47-11.25 |
| Zhang et al., 2020[16] | 2020.01-2020.03 | China | Ancestral lineage | n: 46 Median: 10 IQR: 7-15 | 10.71 | 8.95-12.47 |
| Yu et al., 2020[122] | 2019.12-2020.02 | China | Ancestral lineage | Median: 7.2 95%CI: 6.4-7.9 | / | / |
| Guo et al., 2020[17] | 2020.01-2020.03 | China | Ancestral lineage | n: 85 Median: 9 IQR: 6-13 | 9.35 | 8.23-10.47 |
| Nie et al., 2020[18] | 2020.01-2020.02 | China | Ancestral lineage | n: 2907 Median: 5 IQR: 2.0-8.0 | 5.00 | 4.84-5.16 |
| Liu et al., 2021 [123] | 2020.01-2021.02 | China | Ancestral lineage | Median: 71.5 95%CI: 38.0-134.6 | / | / |
| Wang et al., 2020[124] | 2020.01-2020.02 | China | Ancestral lineage | Mean: 7.4 Median: 7 | / | / |
| Du et al., 2021[21] | 2020.03-2020.07 | China | Ancestral lineage | n: 75 Median: 8.5 IQR: 6.0-12.0 | 8.85 | 7.83-9.88 |
| Lai et al., 2020[125] | 2020.01-2020.03 | China | Ancestral lineage | Median: 4.2 95%CI: 4.0-4.5 | / | / |
| Jia et al., 2020[126] | 2020.01-2020.02 | China | Ancestral lineage | Mean: 6.28 Range:1.0-14.0 | / | / |
| Hua et al., 2020[24] | 2019.12-2020.02 | China | Ancestral lineage | n: 43 Mean: 9.1 SD: 3.7 | 9.10 | 7.99-10.21 |
| Wong et al., 2020[25] | 2020.03-2020.04 | Brunei Darussalam | Ancestral lineage | n: 15 Median: 5 Range: 1.0-11.0 | 5.34 | 3.89-6.80 |
| Zhang et al., 2021[127] | 2020.01-2020.01 | China | Ancestral lineage | Mean: 4 | / | / |
| Hu et al., 2021[29] | 2020.01-2020.04 | China | Ancestral lineage | n: 268 Median: 5.7 IQR: 3.2-8.8 | 5.91 | 5.41-6.41 |
| Zhao et al., 2021[30] | 2019.12-2020.04 | China | Ancestral lineage | Mean: 6.8 95%CI: 6.2-7.5 | 6.80 | 6.15-7.45 |
| Lau et al., 2021[31] | 2020.01-2020.01 | China | Ancestral lineage | Mean: 4.8 95%CI: 4.1-5.6 | 4.80 | 4.05-5.55 |
| Deng et al., 2021[32] | 2020.01-2020.01 | China | Ancestral lineage | Mean: 9.1 95%CI: 7.86-9.66 | 9.10 | 8.20-10.00 |
| Böhm et al., 2021[33] | 2020.01-2020.03 | Germany | Ancestral lineage | n: 256 Mean: 4.6 SD: 3 | 4.60 | 4.23-4.97 |
| Böhmer et al., 2020[34] | 2020.01-2020.02 | Germany | Ancestral lineage | n: 12 Median: 4 IQR: 2.3-4.3 | 3.49 | 2.55-4.43 |
| Wu et al., 2020[128] | 2020.01-2020.02 | China | Ancestral lineage | Median: 4.3 95%CI: 3.4-5.3 | / | / |
| Yang et al., 2020[35] | 2020.01-2020.02 | China | Ancestral lineage | n: 178 Median: 5.4 Range: 1.0-21.0 | 5.82 | 5.28-6.37 |
| Zhang et al., 2020[36] | 2019.12-2020.02 | China | Ancestral lineage | n: 49 Mean: 5.2 SD: 2.59 | 5.20 | 4.47-5.93 |
| Backer et al., 2020[37] | 2020.01-2020.01 | China | Ancestral lineage | Mean: 6.4 95%CrI: 5.6-7.7 | 6.40 | 5.35-7.45 |
| Bui et al., 2020[38] | 2020.01-2020.04 | Vietnam | Ancestral lineage | Mean: 6.4 95%CrI: 4.89-8.5 | 6.40 | 4.60-8.21 |
| Cheng et al., 2021[39] | 2020.01-2020.09 | China | Ancestral lineage | Mean: 7.1 95%CI: 7-7.2 | 7.10 | 7.00-7.20 |
| Han,2020[40] | 2020.03-2020.03 | South Korea | Ancestral lineage | n: 8 Median: 5.5 Range: 3-12 | 6.41 | 4.22-8.60 |
| Kong,2020[41] | 2020.01-2020.02 | China | Ancestral lineage | Mean: 8.5 95%CI: 7.8-9.2 | 8.50 | 7.80-9.20 |
| Men et al., 2020[129] | 2019.12-2020.02 | China | Ancestral lineage | Mean: 5.84 Median: 5 | / | / |
| Xiao et al., 2021[42] | 2019.12-2020.02 | China | Ancestral lineage | n: 217 Mean: 8.58 SD: 4.65 | 8.58 | 7.96-9.20 |
| Xiao et al., 2020[43] | 2019.12-2020.02 | China | Ancestral lineage | Mean: 8.98 95%CI: 7.98-9.9 | 8.98 | 8.02-9.94 |
| Ratovoson et al., 2022[44] | 2020.03-2020.07 | Madagascar | Ancestral lineage | Mean: 4.1 95%CI: 0.7-7.5 | 4.10 | 0.70-7.50 |
| Bi et al., 2020[46] | 2020.01-2020.02 | China | Ancestral lineage | Median: 4.8 95%CI: 4.2-5.4 | / | / |
| Chong et al., 2021[130] | 2020.03-2020.04 | Malaysia | Ancestral lineage | Median: 4.7 95%CI: 3.5-6.4 | / | / |
| Dai et al., 2020[52] | 2020.01-2020.02 | China | Ancestral lineage | n: 180 Mean: 5.83 SD: 3.73 | 5.83 | 5.29-6.37 |
| Huang et al., 2020[53] | 2020.01-2020.02 | China | Ancestral lineage | n: 6 Median: 2 Range: 1-4 | 2.26 | 1.31-3.20 |
| Li et al., 2020[131] | 2019.12-2020.02 | China | Ancestral lineage | Mean: 7.2 SD: 4.11 | / | / |
| Areekal et al., 2021[54] | 2020.06-2020.07 | India | Ancestral lineage | Mean: 4.22 95%CI: 3.71-4.65 | 4.22 | 3.75-4.69 |
| Cao et al., 2020[132] | 2020.01-2020.02 | China | Ancestral lineage | Median: 5.08 95%CI: 4.17-6.21 | / | / |
| Chun et al., 2020[133] | 2020.01-2020.03 | South Korea | Ancestral lineage | Median: 2.87 95%CI: 2.33-3.50 | / | / |
| Tindale et al., 2020[55] | 2020.01-2020.02 | Singapore | Ancestral lineage | Mean: 4.91 95%CI: 4.35-5.69 | 4.91 | 4.24-5.58 |
| Tindale et al., 2020[55] | 2020.01-2020.02 | China | Ancestral lineage | Mean: 7.54 95%CI: 6.76-8.56 | 7.54 | 6.64-8.44 |
| Won et al., 2021[56] | 2020.01-2020.02 | South Korea | Ancestral lineage | Mean: 5.53 95%CI: 3.98-8.09 | 5.53 | 3.48-7.58 |
| Xia et al., 2020[57] | 2020.12-2020.01 | China | Ancestral lineage | Mean: 4.9 95%CI: 4.4-5.4 | 4.90 | 4.40-5.40 |
| Zhao et al., 2021[58] | 2020.01-2020.05 | China | Ancestral lineage | Mean: 6.5 95%CI: 5.6-7.4 | 6.50 | 5.60-7.40 |
| Lauer et al., 2020[134] | 2020.01-2020.02 | China | Ancestral lineage | Median: 5.1 95%CI: 4.5-5.8 | / | / |
| Emecen et al., 2021[59] | 2020.03-2020.11 | Turkey | Ancestral lineage | Median: 3.99 95%CI: 3.25-4.84 | / | / |
| Li et al., 2020[60] | 2020.01-2020.02 | China | Ancestral lineage | Mean: 8.67 95%CI: 8.34-9.02 | 8.67 | 8.33-9.01 |
| Bernal Lopez et al., 2022[62] | 2020.01-2020.03 | UK | Ancestral lineage | n: 45 Mean: 4.51 SD: 2.66 | 4.51 | 3.73-5.29 |
| Bernal Lopez et al., 2022[62] | 2020.01-2020.03 | UK | Ancestral lineage | n: 12 Mean: 4.75 SD: 2.34 | 4.75 | 3.43-6.07 |
| Qin et al., 2020[73] | 2019.12-2020.02 | China | Ancestral lineage | Mean: 8.29 95%CrI: 7.67-8.90 | 8.29 | 7.67-8.90 |
| Ren et al., 2021[74] | 2020.12-2020.01 | China | Ancestral lineage | Mean: 5.3 95%CI: 4.6-6 | 5.30 | 4.60-6.00 |
| Achangwa et al., 2022[135] | 2020.01-2020.10 | South Korea | Ancestral lineage | Median: 4.6 95%CI: 3.9-4.9 | / | / |
| Zhao, et al., 2023[136] | 2020.01-2020.02 | China | Ancestral lineage | n: 97 Median: 10 | / | / |
| Zhao, et al., 2023[136] | 2020.04-2020.05 | China | Ancestral lineage | n: 46 Median: 8 | / | / |
| Zhao, et al., 2023[136] | 2021.01-2021.02 | China | Ancestral lineage | n: 435 Median: 5 | / | / |
| Hammond, et al., 2022[83] | 2020.03-2020.04 | New Zealand | Ancestral lineage | Mean: 3.4 95%CI: 2.7-4.2 | 3.40 | 2.65-4.15 |
| Manica et al., 2022[86] | 2021.03-2021.04 | Italy | Alpha variant | Mean: 4.9 95%CrI: 4.4-5.4 | 4.90 | 4.40-5.40 |
| Tanaka et al., 2022[87] | 2021.04-2021.05 | Japan | Alpha variant | n: 51 Mean: 4.94 SD: 2.19 | 4.94 | 4.34-5.54 |
| Investigation team, 2021[89] | 2020.12-2021.01 | France | Beta variant | n: 10 Median: 4.5 IQR: 2-7 | 4.50 | 1.93-7.07 |
| Manica et al., 2022[86] | 2021.08-2021.10 | Italy | Delta variant | Mean: 4.5 95%CrI: 4.0-5.0 | 4.50 | 4.00-5.00 |
| Li et al., 2022[90] | 2021.05-2021.06 | China | Delta variant | Mean: 6.5 95%CI: 5.86-7.2 | 6.50 | 5.83-7.17 |
| Ma et al., 2022[137] | 2021.05-2021.06 | China | Delta variant | Mean: 5 IQR: 3.0-7.0 | / | / |
| Del Águila-Mejía et al., 2022[138] | 2021.12-2021.12 | Spain | Delta variant | Mean: 3.3 SD: 2.7 | / | / |
| Homma et al., 2021[139] | 2021.03-2021.03 | Japan | Delta variant | Mean: 3.53 Median: 3 | / | / |
| Kang et al., 2022[91] | 2021.05-2021.06 | China | Delta variant | Mean: 5.8 95%CrI: 5.1-6.5 | 5.80 | 5.10-6.50 |
| Ogata et al., 2022[92] | 2020.08-2021.09 | Japan | Delta variant | Mean: 3.7 95%CrI: 3.4-4.0 | 3.70 | 3.40-4.00 |
| Backer et al., 2022[96] | 2021.12-2022.01 | Netherlands | Delta variant | Mean: 4.4 95%CrI: 4.0-4.8 | 4.40 | 4.00-4.80 |
| Li et al., 2022[98] | 2021.08-2021.08 | China | Delta variant | Mean: 4 95%CI: 2.0-4.8 | 4.00 | 2.60-5.40 |
| McAleavey et al., 2022[100] | 2021.11-2021.11 | Ireland | Delta variant | n: 171 Mean: 4.1 SD: 2.4 | 4.10 | 3.74-4.46 |
| Sang Woo Park, 2022[101] | 2021.12-2021.12 | Netherlands | Delta variant | Mean: 4.1 95%CI: 3.8-4.4 | 4.10 | 3.80-4.40 |
| Zhang et al., 2021[102] | 2021.05-2021.06 | China | Delta variant | Mean: 4.4 95%CI: 3.9-5.0 | 4.40 | 3.85-4.95 |
| Liu et al., 2022[103] | 2021.06-2021.06 | South Korea | Delta variant | Mean: 6.5 95%CI: 5.3-7.7 | 6.50 | 5.30-7.70 |
| Deng et al., 2022[140] | 2021.09-2021.09 | China | Delta variant | Median: 4 | / | / |
| Zeng, et al., 2023[105] | 2021.04-2021.07 | Singapore | Delta variant | Mean: 4.3 95%CI: 1.3-10.1 | 4.30 | -0.10-8.70 |
| Yan, et al., 2022[141] | 2021.10-2021.11 | China | Delta variant | n: 5 Mean: 4 | / | / |
| Wright, et al., 2022[142] | 2021.05-2021.12 | Australia | Delta variant | n: 16 Mean: 4.3 | / | / |
| Ogata, et al., 2023[107] | 2021.07-2021.09 | Japan | Delta variant | Mean: 3.7 95%CI: 3.4-4.0 | 3.70 | 3.40-4.00 |
| Luo, et al., 2023[108] | 2021.07-2021.08 | China | Delta variant | n: 71 Mean: 5.3 SD: 3.35 | 5.30 | 4.52-6.08 |
| Guo, et al., 2023[109] | 2022.01-2022.02 | China | Delta variant | Mean: 3.8 95%CrI: 3.1-4.8 | 3.80 | 2.95-4.65 |
| Manica et al., 2022[111] | 2022.01-2022.01 | Italy | Omicron variant | Mean: 3.49 95%CrI: 3.19-3.77 | 3.49 | 3.20-3.78 |
| Mefsin et al., 2022[112] | 2021.12-2022.01 | China | Omicron variant | n: 57 Mean: 4.58 SD: 1.72 | 4.58 | 4.13-5.03 |
| Mefsin et al., 2022[112] | 2021.12-2022.01 | China | Omicron variant | n: 23 Mean: 4.42 SD: 1.42 | 4.42 | 3.84-5.00 |
| Del Águila-Mejía et al., 2022[138] | 2021.12-2021.12 | Spain | Omicron variant | Mean: 3.1 SD: 2.6 | / | / |
| Brandal et al., 2021[113] | 2021.11-2021.12 | Norway | Omicron variant | n: 81 Median: 3 IQR: 3.0-4.0 | 3.35 | 3.19-3.52 |
| Backer et al., 2022[96] | 2021.12-2022.01 | Netherlands | Omicron variant | Mean: 3.2 95%CrI: 2.9-3.6 | 3.20 | 2.85-3.55 |
| Tanaka et al., 2022[87] | 2022.01-2022.01 | Japan | Omicron variant | n: 77 Mean: 3.03 SD: 1.33 | 3.03 | 2.73-3.33 |
| Sang Woo Park, 2022[101] | 2021.12-2021.12 | Netherlands | Omicron variant | Mean: 4.2 95%CI: 3.6-4.9 | 4.20 | 3.55-4.85 |
| HelmSDal et al., 2022[116] | 2021.12-2021.12 | Faroe Islands | Omicron variant | Mean: 3.24 95%CI: 2.87-3.6 | 3.24 | 2.88-3.61 |
| Liu et al., 2022[103] | 2021.11-2021.12 | South Korea | Omicron variant | Mean: 3.5 95%CI: 2.5-3.8 | 3.50 | 2.85-4.15 |
| Zeng, et al., 2023[105] | 2021.12-2023.03 | Singapore | Omicron variant | Mean: 2.8 95%CI: 0.8-7.0 | 2.80 | -0.30-5.90 |
| Xiong, et al., 2023[117] | 2022.06-2022.06 | China | Omicron variant | n: 500 Mean: 3.27 SD: 1.05 | 3.27 | 3.18-3.36 |
| Wei, et al., 2023[118] | 2022.04-2022.04 | China | Omicron variant | n: 52 Mean: 4.6 SD: 2.1 | 4.60 | 4.03-5.17 |
| Wang, et al., 2023[119] | 2022.08-2022.09 | China | Omicron variant | Mean: 5.7 95%CI: 4.8-6.6 | 5.70 | 4.80-6.60 |
| Ogata, et al., 2023[107] | 2022.07-2022.08 | Japan | Omicron variant | Mean: 2.6 95%CI: 2.5-2.8 | 2.60 | 2.45-2.75 |
| Ogata, et al., 2023[107] | 2022.01-2022.02 | Japan | Omicron variant | Mean: 2.9 95%CI: 2.6-3.2 | 2.90 | 2.60-3.20 |
| Guo, et al., 2023[109] | 2022.01-2022.02 | China | Omicron variant | Mean: 3.4 95%CrI: 2.9-4.0 | 3.40 | 2.85-3.95 |

* The detailed method for calculating the data is described in the Methods.

Abbreviations: SD, standard deviation; CI, confidence interval; CrI, credible interval; IQR, interquartile range.

### Table S5. Characteristic of included articles for realized serial interval and parameters extracted.

| Author | Study period | Location | Strain type | Data extracted | Data for analysis* | |
| --- | --- | --- | --- | --- | --- | --- |
|  |  |  |  |  | Mean | 95%CI |
| Vazirinejad et al., 2020[2] | 2020.03-2020.04 | Iran | Ancestral lineage | Mean: 6.4 95%CI: 5.2-7.6 | 6.40 | 5.20-7.60 |
| Bender et al., 2021[3] | 2020.02-2020.03 | Germany | Ancestral lineage | Median: 3 IQR: 1.0-6.0 | / | / |
| Liu et al., 2020[4] | 2020.01-2020.04 | China | Ancestral lineage | n: 31 Median: 4 Range: -3-24 | 5.52 | 3.21-7.83 |
| Haddad et al., 2021[5] | 2020.02-2020.06 | Lebanon | Ancestral lineage | Mean: 5.24 95%CI: 4.37-6.03 | 5.24 | 4.41-6.07 |
| Shi et al., 2020[6] | 2020.01-2020.03 | China | Ancestral lineage | Mean: 6.31 95%CI: 5.12-7.50 | 6.31 | 5.12-7.50 |
| Gupta et al., 2020[8] | 2020.03-2020.07 | India | Ancestral lineage | Mean: 5.4 95%CI: 4.4-6.4 | 5.40 | 4.40-6.40 |
| Bao et al., 2021[9] | 2020.01-2020.02 | China | Ancestral lineage | Mean: 4.4 95%CI: 3.3-5.4 | 4.40 | 3.35-5.45 |
| Du et al., 2020[10] | 2020.01-2020.02 | China | Ancestral lineage | Mean: 3.96 95%CI: 3.53-4.39 | 3.96 | 3.53-4.39 |
| Li et al., 2020[11] | 2019.12-2020.01 | China | Ancestral lineage | Mean: 7.5 95%CI: 5.3-19 | 7.50 | 0.65-14.35 |
| Zhu et al., 2021[12] | 2021.01-2021.02 | China | Ancestral lineage | Mean: 6.6 95%CI: 0.6-20.0 | 6.60 | -3.10-16.30 |
| Ping et al., 2021[13] | 2019.12-2020.03 | China | Ancestral lineage | n: 52 Mean: 6.14 SD: 2.21 | 6.14 | 5.54-6.74 |
| Ki,2020[14] | 2020.01-2020.01 | South Korea | Ancestral lineage | n: 12 Median: 4 Range: 3-15 | 5.91 | 3.83-8.00 |
| Son et al., 2020[19] | 2020.01-2020.03 | South Korea | Ancestral lineage | Mean: 5.54 95%CI: 4.08-7.01 | 5.54 | 4.08-7.00 |
| Baskaradoss et al., 2021[20] | 2020.02-2020.05 | Kuwait | Ancestral lineage | n: 216 Mean: 3.89 SD: 3.69 | 3.89 | 3.40-4.38 |
| Bae et al., 2020[143] | 2020.02-2020.03 | South Korea | Ancestral lineage | Mean: 5.2 SD: 3.8 | / | / |
| Thway et al., 2020[22] | 2020.03-2020.07 | Myanmar | Ancestral lineage | n: 169 Median: 4 IQR: 2-5 | 3.65 | 3.31-3.99 |
| Kwok et al., 2020[23] | 2020.01-2020.02 | China | Ancestral lineage | Mean: 4.77 95%CrI: 3.47-6.90 | 4.77 | 3.05-6.48 |
| Wong et al., 2020[25] | 2020.03-2020.04 | Brunei Darussalam | Ancestral lineage | Mean: 5.4 95%CI: 4.3-6.5 | 5.40 | 4.30-6.50 |
| Haw et al., 2020[26] | 2020.04-2020.05 | Philippines | Ancestral lineage | Mean: 6.9 95%CI: 5.81-8.41 | 6.90 | 5.60-8.20 |
| Hu et al., 2021[29] | 2020.01-2020.04 | China | Ancestral lineage | n: 245 Median: 4.8 IQR: 0.8-9.4 | 5.01 | 4.21-5.81 |
| Böhm et al., 2021[33] | 2020.01-2020.03 | Germany | Ancestral lineage | n: 53 Mean: 3.9 SD: 2.2 | 3.90 | 3.31-4.49 |
| Böhmer et al., 2020[34] | 2020.01-2020.02 | Germany | Ancestral lineage | Median: 4 IQR: 3-5 | / | / |
| Wu et al., 2020[128] | 2020.01-2020.02 | China | Ancestral lineage | Median: 5.1 95%CI: 4.3-6.2 | / | / |
| Yang et al., 2020[35] | 2020.01-2020.02 | China | Ancestral lineage | n: 131 Median: 4.6 Range: -4-13 | 4.59 | 4.03-5.15 |
| Zhang et al., 2020[36] | 2019.12-2020.02 | China | Ancestral lineage | Mean: 5.1 95%CI: 1.3-11.6 | 5.10 | -0.05-10.25 |
| Ratovoson et al., 2022[44] | 2020.03-2020.07 | Madagascar | Ancestral lineage | Mean: 6 95%CI: 2.4-9.6 | 6.00 | 2.40-9.60 |
| Geismar et al., 2021[80] | 2020.09-2021.01 | UK | Ancestral lineage | Mean: 2.7 95%CI: 1.5-4 | 2.70 | 1.45-3.95 |
| Aghaali et al., 2020[45] | 2020.02-2020.03 | Iran | Ancestral lineage | n: 37 Mean: 4.55 SD: 3.3 | 4.55 | 3.49-5.61 |
| Bi et al., 2020[46] | 2020.01-2020.02 | China | Ancestral lineage | Mean: 6.3 95%CI: 5.2-7.6 | 6.30 | 5.10-7.50 |
| Chu et al., 2021[144] | 2020.07-2020.08 | USA | Ancestral lineage | Median: 5 95%CI: 4.0-6.5 | / | / |
| Expert Taskforce,2020[47] | 2019.12-2020.03 | Japan | Ancestral lineage | n: 26 Median: 2 IQR: 2-4 | 2.71 | 2.12-3.31 |
| McAloon et al., 2021[48] | 2020.04-2020.12 | Ireland | Ancestral lineage | Mean: 4 95%CI: 3.7-4.3 | 4.00 | 3.70-4.30 |
| Ng et al., 2021[145] | 2020.01-2020.04 | Singapore | Ancestral lineage | Median: 3.28 95%CrI: −5.41-13.11 | / | / |
| Talmoudi et al., 2020[49] | 2020.03-2020.05 | Tunisia | Ancestral lineage | Mean: 5.3 95%CI: 4.66-5.95 | 5.30 | 4.65-5.95 |
| Wang et al., 2020[50] | 2020.01-2020.02 | China | Ancestral lineage | Mean: 5.9 95%CI: 3.9-9.6 | 5.90 | 3.05-8.75 |
| Kwok et al., 2021[51] | 2020.01-2020.08 | China | Ancestral lineage | n: 558 Mean: 4.74 SD: 4.24 | 4.74 | 4.39-5.09 |
| Chong et al., 2021[130] | 2020.03-2020.04 | Malaysia | Ancestral lineage | Median: 5.3 95%CI: 4.3-6.5 | / | / |
| Huang et al., 2020[53] | 2020.01-2020.02 | China | Ancestral lineage | n: 7 Median: 1 Range: 0-4 | 1.48 | 0.39-2.58 |
| Areekal et al., 2021[54] | 2020.06-2020.07 | India | Ancestral lineage | Mean: 5.24 95%CI: 4.764-5.716 | 5.24 | 4.76-5.72 |
| Cao et al., 2020[132] | 2020.01-2020.02 | China | Ancestral lineage | Median: 6 95%CI: 5-7 | / | / |
| Chun et al., 2020[133] | 2020.01-2020.03 | South Korea | Ancestral lineage | Median: 3.56 95%CI: 2.72-4.44 | / | / |
| Tindale et al., 2020[55] | 2020.01-2020.02 | China | Ancestral lineage | Mean: 4.31 95%CI: 2.91-5.72 | 4.31 | 2.90-5.71 |
| Tindale et al., 2020[55] | 2020.01-2020.02 | Singapore | Ancestral lineage | Mean: 4.17 95%CI: 2.44-5.89 | 4.17 | 2.45-5.89 |
| Won et al., 2021[56] | 2020.01-2020.02 | South Korea | Ancestral lineage | Mean: 6.45 95%CI: 4.32-9.65 | 6.45 | 3.79-9.12 |
| Xia et al., 2020[57] | 2020.12-2020.01 | China | Ancestral lineage | n: 74 Mean: 4.1 SD: 3.3 | 4.10 | 3.35-4.85 |
| Zhao et al., 2021[58] | 2020.01-2020.05 | China | Ancestral lineage | Mean: 5.95 95%CI: 5.24-6.66 | 5.95 | 5.24-6.66 |
| Emecen et al., 2021[59] | 2020.03-2020.11 | Turkey | Ancestral lineage | n: 36 Mean: 4.81 SD: 3.24 | 4.81 | 3.75-5.87 |
| Li et al., 2020[60] | 2020.01-2020.02 | China | Ancestral lineage | Mean: 6.05 95%CI: 5.68-6.44 | 6.05 | 5.67-6.43 |
| Ali et al., 2020[61] | 2020.01-2020.02 | China | Ancestral lineage | Mean: 5.1 95%CrI: 4.7-5.5 | 5.10 | 4.70-5.50 |
| Bernal Lopez et al., 2022[62] | 2020.01-2020.03 | UK | Ancestral lineage | Mean: 4.67 | / | / |
| Cereda et al., 2021[63] | 2020.01-2020.03 | Italy | Ancestral lineage | Mean: 6.6 95%CrI: 0.7-19 | 6.60 | -2.55-15.75 |
| Hong et al., 2020[64] | 2020.01-2020.08 | South Korea | Ancestral lineage | n: 1567 Mean: 4.02 SD: 4.91 | 4.02 | 3.78-4.26 |
| Lavezzo et al., 2020[65] | 2020.02-2020.03 | Italy | Ancestral lineage | Mean: 7.2 95%CI: 5.9-9.6 | 7.20 | 5.35-9.05 |
| Liu et al., 2020[66] | 2020.01-2020.03 | China | Ancestral lineage | n: 116 Mean: 5.81 SD: 3.24 | 5.81 | 5.22-6.40 |
| Najafi et al., 2020[67] | 2020.02-2020.03 | Iran | Ancestral lineage | n: 21 Mean: 5.71 SD: 3.89 | 5.71 | 4.05-7.37 |
| Prete et al., 2020[68] | 2022.02-2022.03 | Brazil | Ancestral lineage | n: 65 Mean: 2.97 SD: 3.29 | 2.97 | 2.17-3.77 |
| Reed et al., 2021[69] | 2022.03-2020.07 | USA | Ancestral lineage | Mean: 5.68 95%CI: 5.27-6.08 | 5.68 | 5.27-6.08 |
| Ryu et al., 2021[70] | 2020.01-2020.04 | South Korea | Ancestral lineage | Mean: 4 95%CrI: 3.7-4.3 | 4.00 | 3.70-4.30 |
| Ryu et al., 2021[70] | 2020.04-2020.08 | South Korea | Ancestral lineage | Mean: 3.2 95%CrI: 3.0-3.5 | 3.20 | 2.95-3.45 |
| Saurabh et al., 2020[71] | 2020.03-2020.07 | India | Ancestral lineage | n: 103 Mean: 6.23 SD: 3.49 | 6.23 | 5.56-6.90 |
| Thai et al., 2021[72] | 2020.01-2020.05 | Vietnam | Ancestral lineage | Mean: 3.24 95%CI: 1.38-5.10 | 3.24 | 1.38-5.10 |
| Wang et al., 2021[146] | 2020.02-2020.07 | USA | Ancestral lineage | Mean: 4.99 | / | / |
| Ren et al., 2021[74] | 2019.12-2020.01 | China | Ancestral lineage | Mean: 5.7 95%CI: 4.7-6.8 | 5.70 | 4.65-6.75 |
| Zhao et al., 2020[75] | 2020.01-2020.02 | China | Ancestral lineage | Mean: 4.9 95%CI: 3.6-6.2 | 4.90 | 3.60-6.20 |
| Adam et al., 2020[76] | 2020.01-2020.04 | China | Ancestral lineage | n: 142 Mean: 5.8 SD: 4.43 | 5.80 | 5.07-6.53 |
| Buchholz et al., 2022[81] | 2020.03-2020.11 | Germany | Ancestral lineage | Mean: 4.8 95%CI: 4.74-4.85 | 4.80 | 4.75-4.85 |
| Althobaity et al., 2022[78] | 2020.03-2020.04 | Saudi Arabia | Ancestral lineage | Mean: 5.1 95%CI: 5.0-5.5 | 5.10 | 4.85-5.35 |
| Wang et al., 2022[79] | 2020.06-2020.07 | China | Ancestral lineage | n: 34 Median: 3.2 IQR: 0.7-5.8 | 3.24 | 1.91-4.56 |
| Somda, et al., 2022[82] | 2020.03-2020.05 | Burkina Faso | Ancestral lineage | n: 113 Mean: 5.92 SD: 5.8 | 5.92 | 4.85-6.99 |
| Hammond, et al., 2022[83] | 2020.03-2020.04 | New Zealand | Ancestral lineage | Mean: 4 95%CI: 3.2-4.7 | 4.00 | 3.25-4.75 |
| Geismar, et al., 2022[84] | 2020.09-2022.08 | UK | Ancestral lineage | Mean: 2.29 95%CrI: 1.39-2.94 | 2.29 | 1.52-3.06 |
| Manica et al., 2022[86] | 2021.03-2021.04 | Italy | Alpha variant | Mean: 2.4 95%CrI: 2.20-2.60 | 2.40 | 2.20-2.60 |
| Geismar et al., 2021[80] | 2020.09-2021.01 | UK | Alpha variant | Mean: 3.6 95%CI: 2.5-4.7 | 3.60 | 2.50-4.70 |
| Buchholz et al., 2022[81] | 2021.03-2021.05 | Germany | Alpha variant | Mean: 4.5 95%CI: 4.46-4.54 | 4.50 | 4.46-4.54 |
| Geismar, et al., 2022[84] | 2020.09-2022.08 | UK | Alpha variant | Mean: 3.37 95%CrI: 2.52-4.04 | 3.37 | 2.61-4.13 |
| Manica et al., 2022[86] | 2021.08-2021.10 | Italy | Delta variant | Mean: 2.56 95%CrI: 2.37-2.74 | 2.56 | 2.38-2.75 |
| Li et al., 2022[90] | 2021.05-2021.06 | China | Delta variant | Mean: 4.24 95%CI: 3.35-5.14 | 4.24 | 3.35-5.13 |
| Ma et al., 2022[137] | 2021.05-2021.06 | China | Delta variant | Median: 3 IQR: 1.0-5.0 | / | / |
| Del Águila-Mejía et al., 2022[138] | 2021.12-2021.12 | Spain | Delta variant | Mean: 5.4 SD: 3.1 | / | / |
| Kang et al., 2022[91] | 2021.05-2021.06 | China | Delta variant | Mean: 6.7 95%CrI: 4.9-9.3 | 6.70 | 4.50-8.90 |
| Ogata et al., 2022[92] | 2020.08-2021.09 | Japan | Delta variant | Mean: 2.8 | / | / |
| Pung et al., 2021[93] | 2021.04-2021.05 | Singapore | Delta variant | n: 32 Mean: 3.3 SD: 3.08 | 3.30 | 2.23-4.37 |
| Ryu et al., 2022[94] | 2021.07-2021.09 | South Korea | Delta variant | Mean: 3.6 95%CrI: 3.5-3.6 | 3.60 | 3.55-3.65 |
| Kremer et al., 2022[95] | 2021.11-2021.12 | Belgium | Delta variant | Mean: 3 95%CrI: 2.73-3.26 | 3.00 | 2.74-3.26 |
| Backer et al., 2022[96] | 2021.12-2021.12 | Netherlands | Delta variant | n: 761 Mean: 4.1 SD: 2.8 | 4.10 | 3.90-4.30 |
| Backer et al., 2022[96] | 2021.12-2021.12 | Netherlands | Delta variant | n: 158 Mean: 3.5 SD: 2.8 | 3.50 | 3.06-3.94 |
| Backer et al., 2022[96] | 2021.12-2021.12 | Netherlands | Delta variant | n: 572 Mean: 3.2 SD: 2.6 | 3.20 | 2.99-3.41 |
| Backer et al., 2022[96] | 2021.12-2021.12 | Netherlands | Delta variant | n: 130 Mean: 2.94 SD: 2.16 | 2.94 | 2.57-3.31 |
| Hwang et al., 2022[97] | 2021.06-2021.07 | South Korea | Delta variant | Mean: 3.26 95%CrI: 2.92-3.60 | 3.26 | 2.92-3.60 |
| Li et al., 2022[98] | 2021.08-2021.08 | China | Delta variant | Mean: 2.6 95%CI: 1.9-3.6 | 2.60 | 1.75-3.45 |
| Song et al., 2022[99] | 2021.07-2021.12 | South Korea | Delta variant | Mean: 3.15 95%CI: 3.10-3.20 | 3.15 | 3.10-3.20 |
| Sang Woo Park,2022[101] | 2021.12-2021.12 | Netherlands | Delta variant | Mean: 3.7 95%CI: 3.5-3.8 | 3.70 | 3.55-3.85 |
| Sang Woo Park,2022[101] | 2021.12-2021.12 | Netherlands | Delta variant | Mean: 3.3 95%CI: 3.0-3.6 | 3.30 | 3.00-3.60 |
| Allen et al., 2022[147] | 2021.12-2021.12 | UK | Delta variant | Median: 4 IQR: 2-6 | / | / |
| Allen et al., 2022[147] | 2021.12-2021.12 | UK | Delta variant | Median: 4 IQR: 2-7 | / | / |
| Zhang et al., 2021[102] | 2021.05-2021.06 | China | Delta variant | Mean: 2.3 95%CI: 1.4-3.3 | 2.30 | 1.35-3.25 |
| Deng et al., 2022[140] | 2021.09-2021.09 | China | Delta variant | Mean: 1.7 SD: 3 | / | / |
| Buchholz et al., 2022[81] | 2021.07-2021.12 | Germany | Delta variant | Mean: 4.19 95%CI: 4.16-4.22 | 4.19 | 4.16-4.22 |
| Ali et al., 2022[104] | 2021.12-2022.02 | China | Delta variant | n: 25 Mean: 4 SD: 2.4 | 4.00 | 3.06-4.94 |
| Zeng, et al., 2023[105] | 2021.04-2021.07 | Singapore | Delta variant | Mean: 4.3 95%CI: 0.9-12.4 | 4.30 | -1.45-10.05 |
| Wang, et al., 2022[106] | 2021.07-2021.08 | China | Delta variant | n: 72 Mean: 4.79 SD: 3.47 | 4.79 | 3.99-5.59 |
| Luo, et al., 2023[108] | 2021.07-2021.08 | China | Delta variant | n: 54 Mean: 4.3 SD: 3.62 | 4.30 | 3.33-5.27 |
| Guo, et al., 2023[109] | 2022.01-2022.02 | China | Delta variant | Mean: 5.8 95%CrI: 4.9-6.7 | 5.80 | 4.90-6.70 |
| Geismar, et al., 2022[84] | 2020.09-2022.08 | UK | Delta variant | Mean: 3.11 95%CrI: 2.28-3.90 | 3.11 | 2.30-3.92 |
| Song et al., 2022[110] | 2021.11-2021.12 | South Korea | Omicron variant | n: 12 Mean: 2.9 SD: 1.6 | 2.90 | 1.99-3.81 |
| Manica et al., 2022[111] | 2022.01-2022.01 | Italy | Omicron variant | Mean: 2.38 95%CrI: 2.30-2.47 | 2.38 | 2.29-2.46 |
| Mefsin et al., 2022[112] | 2021.12-2022.01 | China | Omicron variant | n: 30 Mean: 3.3 SD: 1.95 | 3.30 | 2.60-4.00 |
| Mefsin et al., 2022[112] | 2021.12-2022.01 | China | Omicron variant | n: 13 Mean: 2.72 SD: 1.51 | 2.72 | 1.90-3.54 |
| Del Águila-Mejía et al., 2022[137] | 2021.12-2021.12 | Spain | Omicron variant | Mean: 4.8 SD: 3 | / | / |
| Kremer et al., 2022[95] | 2021.11-2021.12 | Belgium | Omicron variant | Mean: 2.75 95%CrI: 2.65-2.86 | 2.75 | 2.64-2.85 |
| Backer et al., 2022[96] | 2021.12-2021.12 | Netherlands | Omicron variant | n: 164 Mean: 3.5 SD: 2.4 | 3.50 | 3.13-3.87 |
| Backer et al., 2022[96] | 2021.12-2021.12 | Netherlands | Omicron variant | n: 71 Mean: 3.3 SD: 2.4 | 3.30 | 2.74-3.86 |
| Backer et al., 2022[96] | 2021.12-2021.12 | Netherlands | Omicron variant | n: 480 Mean: 3 SD: 2.3 | 3.00 | 2.79-3.21 |
| Backer et al., 2022[96] | 2021.12-2021.12 | Netherlands | Omicron variant | n: 193 Mean: 2.87 SD: 2.07 | 2.87 | 2.58-3.16 |
| Kim et al., 2022[114] | 2021.11-2021.12 | South Korea | Omicron variant | Mean: 3.78 95%CrI: 3.02-4.54 | 3.78 | 3.02-4.54 |
| Shim et al., 2022[115] | 2021.11-2022.01 | South Korea | Omicron variant | Mean: 4.2 95%CI: 3.8-4.5 | 4.20 | 3.85-4.55 |
| Sang Woo Park,2022[101] | 2021.12-2021.12 | Netherlands | Omicron variant | Mean: 3.1 95%CI: 2.9-3.3 | 3.10 | 2.90-3.30 |
| Sang Woo Park,2022[101] | 2021.12-2021.12 | Netherlands | Omicron variant | Mean: 3 95%CI: 2.7-3.3 | 3.00 | 2.70-3.30 |
| Allen et al., 2022[147] | 2021.12-2021.12 | UK | Omicron variant | Median: 3 IQR: 2-5 | / | / |
| Allen et al., 2022[147] | 2021.12-2021.12 | UK | Omicron variant | Median: 3 IQR: 1-5 | / | / |
| Buchholz et al., 2022[81] | 2022.01-2022.04 | Germany | Omicron variant | Mean: 3.61 95%CI: 3.56-3.66 | 3.61 | 3.56-3.66 |
| Ali et al., 2022[104] | 2021.12-2022.02 | China | Omicron variant | n: 30 Mean: 3.3 SD: 2 | 3.30 | 2.58-4.02 |
| Ali et al., 2022[104] | 2021.12-2022.02 | China | Omicron variant | n: 174 Mean: 3.6 SD: 1.8 | 3.60 | 3.33-3.87 |
| Zeng, et al., 2023[105] | 2021.12-2023.03 | Singapore | Omicron variant | Mean: 2.6 95%CI: 0.6-6.8 | 2.60 | -0.50-5.70 |
| Zeng, et al., 2023[105] | 2021.01-2023.03 | Singapore | Omicron variant | Mean: 2.6 95%CI: 1.0-5.1 | 2.60 | 0.55-4.65 |
| Wei, et al., 2023[118] | 2022.04-2022.04 | China | Omicron variant | n: 234 Mean: 3.9 SD: 3.6 | 3.90 | 3.44-4.36 |
| Guo, et al., 2023[120] | 2022.07-2022.07 | China | Omicron variant | Mean: 2.8 95%CrI: 1.5-6.7 | 2.80 | 0.20-5.40 |
| Guo, et al., 2023[120] | 2022.06-2022.07 | China | Omicron variant | Mean: 2.7 95%CrI: 2.1-3.6 | 2.70 | 1.95-3.45 |
| Guo, et al., 2023[120] | 2022.05-2022.06 | China | Omicron variant | Mean: 4.4 95%CrI: 2.6-7.5 | 4.40 | 1.95-6.85 |
| Guo, et al., 2023[109] | 2022.01-2022.02 | China | Omicron variant | Mean: 4.4 95%CrI: 4.1-4.9 | 4.40 | 4.00-4.80 |
| Geismar, et al., 2022[84] | 2020.09-2022.08 | UK | Omicron variant | Mean: 2.02 95%CrI: 1.26-2.84 | 2.02 | 1.23-2.81 |
| Geismar, et al., 2022[84] | 2020.09-2022.08 | UK | Omicron variant | Mean: 2.72 95%CrI: 2.01-3.47 | 2.72 | 1.99-3.45 |
| Geismar, et al., 2022[84] | 2020.09-2022.08 | UK | Omicron variant | Mean: 2.67 95%CrI: 1.90-3.46 | 2.67 | 1.89-3.45 |

* The detailed method for calculating the data is described in the Methods.

Abbreviations: SD, standard deviation; CI, confidence interval; CrI, credible interval; IQR, interquartile range.

### Table S6. Characteristic of included articles for realized generation time and parameters extracted.

| Author | Study period | Location | Strain type | Data extracted | Data for analysis* | |
| --- | --- | --- | --- | --- | --- | --- |
|  |  |  |  |  | Mean | 95%CI |
| Bender et al., 2021[3] | 2020.02-2020.03 | Germany | Ancestral lineage | Median: 3.6 IQR: 1.7-6.6 | / | / |
| Ganyani et al., 2020[27] | 2020.01-2020.02 | China | Ancestral lineage | Mean: 3.95 95%CrI: 3.01-4.91 | 3.95 | 3.00-4.90 |
| Ganyani et al., 2020[27] | 2020.01-2020.02 | Singapore | Ancestral lineage | Mean: 5.2 95%CrI: 3.78-6.78 | 5.20 | 3.70-6.70 |
| Hart et al., 2022[28] | 2020.03-2020.12 | UK | Ancestral lineage | Mean: 4.2 95%CrI: 3.3-5.3 | 4.20 | 3.20-5.20 |
| Hu et al., 2021[29] | 2020.01-2020.04 | China | Ancestral lineage | n: 268 Median: 5.5 IQR: 4.5-6.8 | 5.61 | 5.40-5.81 |
| Zhao et al., 2021[30] | 2019.12-2020.04 | China | Ancestral lineage | Mean: 6.7 95%CI: 5.4-7.6 | 6.70 | 5.60-7.80 |
| Lau et al., 2021[31] | 2020.01-2020.01 | China | Ancestral lineage | Mean: 5.7 95%CI: 4.8-6.5 | 5.70 | 4.85-6.55 |
| Ng et al., 2021[145] | 2020.01-2020.04 | Singapore | Ancestral lineage | Mean: 3.44 95%CI: 2.79-4.11 | 3.44 | 2.78-4.10 |
| Li et al., 2020[131] | 2019.12-2020.02 | China | Ancestral lineage | Mean: 3.3 SD: 1.76 | / | / |
| Li et al., 2020[60] | 2020.01-2020.02 | China | Ancestral lineage | Mean: 4.81 95%CI: 4.13-5.58 | 4.81 | 4.08-5.54 |
| Ferretti et al., 2020[77] | 2019.12-2020.03 | multiple countries | Ancestral lineage | n: 40 Mean: 5 SD: 1.9 | 5.00 | 4.41-5.59 |
| Hart et al., 2022[85] | 2021.02-2021.08 | UK | Alpha variant | Mean: 4.5 95%CrI: 3.7-5.4 | 4.50 | 3.65-5.35 |
| Manica et al., 2022[86] | 2021.03-2021.04 | Italy | Alpha variant | Mean: 4.08 95%CrI: 4.06-4.09 | 4.08 | 4.06-4.10 |
| Amato et al., 2022[88] | 2020.12-2021.02 | Italy | Alpha variant | n: 149 Mean: 4.7 SD: 2.9 | 4.70 | 4.23-5.17 |
| Hart et al., 2022[85] | 2021.02-2021.08 | UK | Delta variant | Mean: 3.2 95%CrI: 2.5-4.2 | 3.20 | 2.35-4.05 |
| Manica et al., 2022[86] | 2021.08-2021.10 | Italy | Delta variant | Mean: 3.96 95%CrI: 3.94-4.01 | 3.96 | 3.92-4.00 |
| Sang Woo Park, 2022[101] | 2021.12-2021.12 | Netherlands | Delta variant | Mean: 3.8 95%CI: 3.7-4.0 | 3.80 | 3.65-3.95 |
| Sang Woo Park, 2022[101] | 2021.12-2021.12 | Netherlands | Delta variant | Mean: 3.5 95%CI: 3.2-3.8 | 3.50 | 3.20-3.80 |
| Zhang et al., 2021[102] | 2021.05-2021.06 | China | Delta variant | Mean: 2.9 95%CI: 2.4-3.3 | 2.90 | 2.45-3.35 |
| Deng et al., 2022[140] | 2021.09-2021.09 | China | Delta variant | Mean: 3.6 SD: 2.6 | / | / |
| Yan, et al., 2022[141] | 2021.10-2021.11 | China | Delta variant | n: 5 Mean: 3 | / | / |
| Luo, et al., 2023[108] | 2021.07-2021.08 | China | Delta variant | n: 71 Mean: 4.4 SD: 2.64 | 4.40 | 3.79-5.01 |
| Manica et al., 2022[111] | 2022.01-2022.01 | Italy | Omicron variant | Mean: 3.59 95%CrI: 3.55-3.60 | 3.59 | 3.56-3.62 |
| Mefsin et al., 2022[112] | 2021.12-2022.01 | China | Omicron variant | Mean: 2.36 95%CI: 2.01-2.77 | 2.36 | 1.98-2.74 |
| Sang Woo Park, 2022[101] | 2021.12-2021.12 | Netherlands | Omicron variant | Mean: 3 95%CI: 2.7-3.2 | 3.00 | 2.75-3.25 |
| Sang Woo Park, 2022[101] | 2021.12-2021.12 | Netherlands | Omicron variant | Mean: 2.9 95%CI: 2.5-3.3 | 2.90 | 2.50-3.30 |
| Wang, et al., 2023[119] | 2022.08-2022.09 | China | Omicron variant | Mean: 2.8 95%CrI: 2.4-3.5 | 2.80 | 2.25-3.35 |

* The detailed method for calculating the data is in the Methods.

Abbreviations: n, sample size; SD, standard deviation; CI, confidence interval; CrI, credible interval; IQR, interquartile range.

### Table S7. Characteristic of included articles for intrinsic generation time and parameters extracted.

| Author | Study period | Location | Strain type | Data extracted | Data for analysis* | |
| --- | --- | --- | --- | --- | --- | --- |
|  |  |  |  |  | Mean | 95%CI |
| Hart et al., 2022[85] | 2021.02-2021.08 | UK | Alpha variant | Mean: 5.5 95%CrI: 4.7-6.5 | 5.50 | 4.60-6.40 |
| Manica et al., 2022[86] | 2021.03-2021.04 | Italy | Alpha variant | Mean: 5.95 95%CrI: 5.57-6.44 | 5.95 | 5.52-6.38 |
| Hart et al., 2022[85] | 2021.02-2021.08 | UK | Delta variant | Mean: 4.7 95%CrI: 4.1-5.6 | 4.70 | 3.95-5.45 |
| Manica et al., 2022[86] | 2021.08-2021.10 | Italy | Delta variant | Mean: 6.62 95%CrI: 6.01-7.31 | 6.62 | 5.97-7.27 |
| Manica et al., 2022[111] | 2022.01-2022.01 | Italy | Omicron variant | Mean: 6.84 95%CrI: 5.72-8.60 | 6.84 | 5.40-8.28 |

* The detailed method for calculating the data is described in the Methods.

Abbreviations: n, sample size; SD, standard deviation; CI, confidence interval; CrI, credible interval; IQR, interquartile range.

### Table S8. Excluded studies and reasons for exclusion.

| **Studies** | **Reason for exclusion** |
| --- | --- |
| [148-161] | Reviews |
| [162-182] | Meta-analyses |
| [183] | Comment |
| [184-187] | Full text unavailable |
| [188] | Study period unavailable |
| [189-193] | Sample size less than 5 |
| [194-198] | Estimate unavailable |
| [199-206] | Data source unknown |
| [207, 208] | Methods not described |
| [209-247] | Unknown epidemiological link |

### **Figure S1. Forest plot for studies of incubation period of COVID-19 caused by different SARS-CoV-2 virus lineages.**

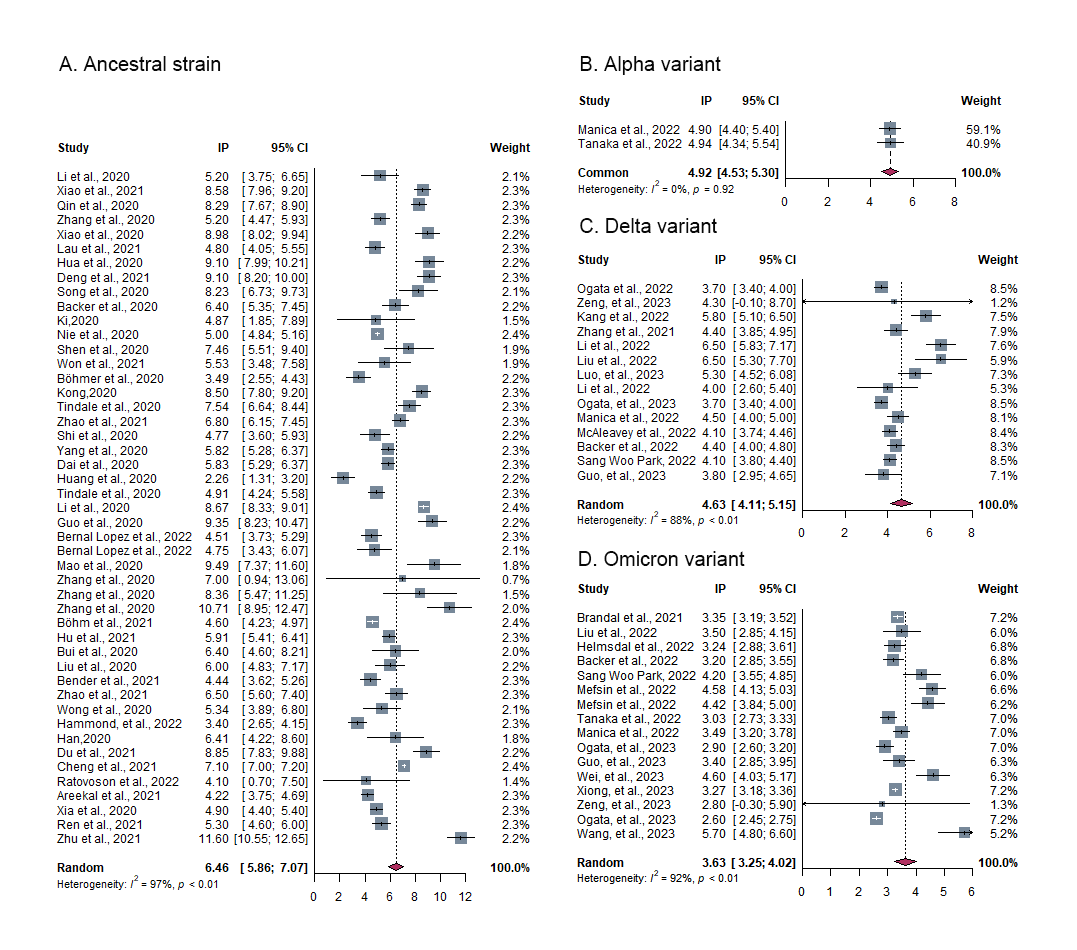

**Figure S1. Forest plot for studies of incubation period of COVID-19 caused by different SARS-CoV-2 virus lineages.** Seventy-nine records were selected and incorporated into meta-analysis. The Beta variant was excluded due to insufficient number of records for a pooled analysis and 29 records were excluded since they did not provide a central or dispersion tendency estimate, nor could one be inferred from the available summary statistics.

### **Figure S2. Forest plot for studies of realized serial interval of COVID-19 caused by different SARS-CoV-2 virus lineages.**

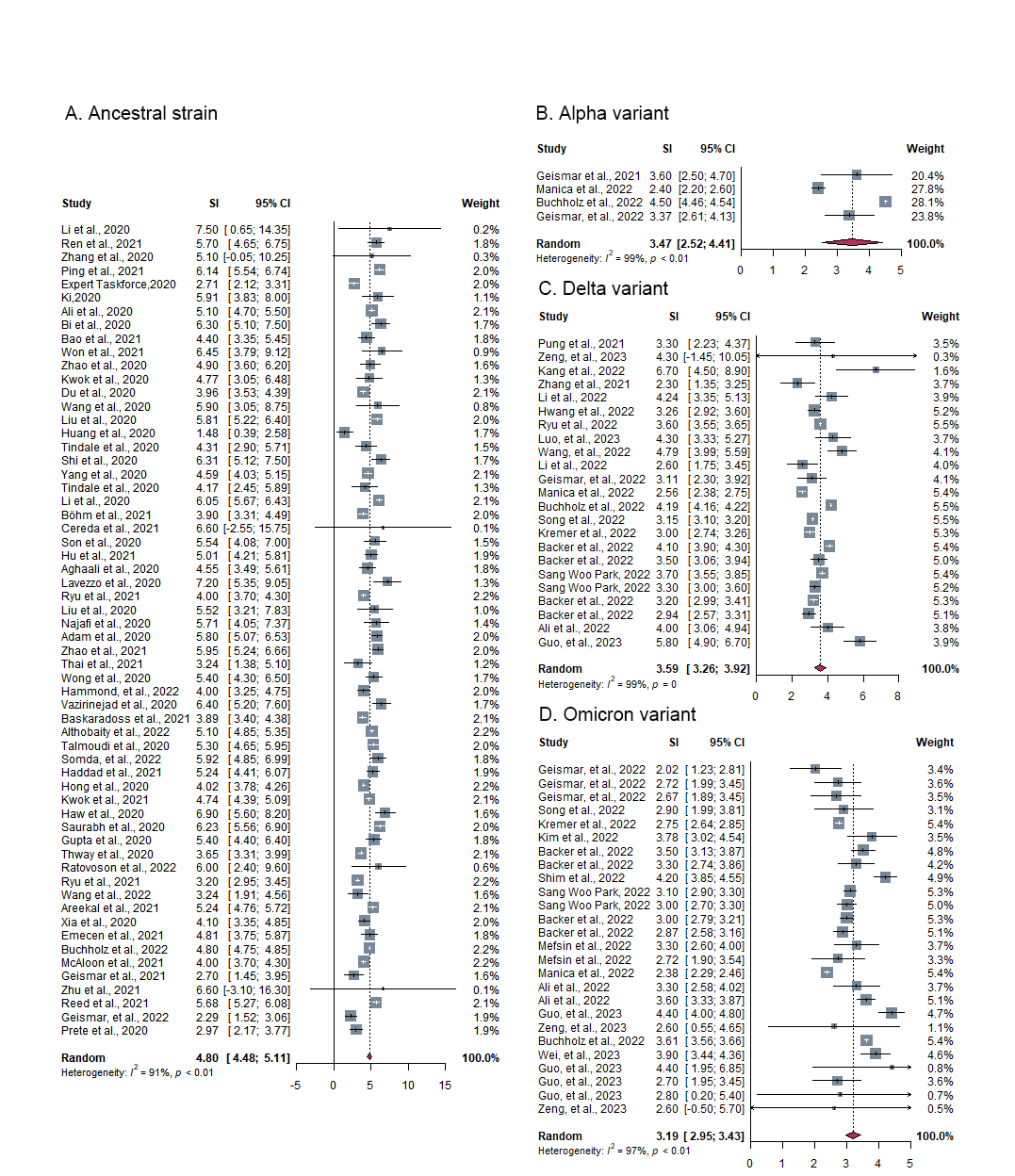

**Figure S2. Forest plot for studies of realized serial interval of COVID-19 caused by different SARS-CoV-2 virus lineages.** One hundred and thirteen records were selected and incorporated into meta-analysis. Twenty records were excluded since they did not provide a central or dispersion tendency estimate, nor could one be inferred from the available summary statistics.

Figure S3. Forest plot for studies of realized generation time of COVID-19 caused by different SARS-CoV-2 virus lineages.

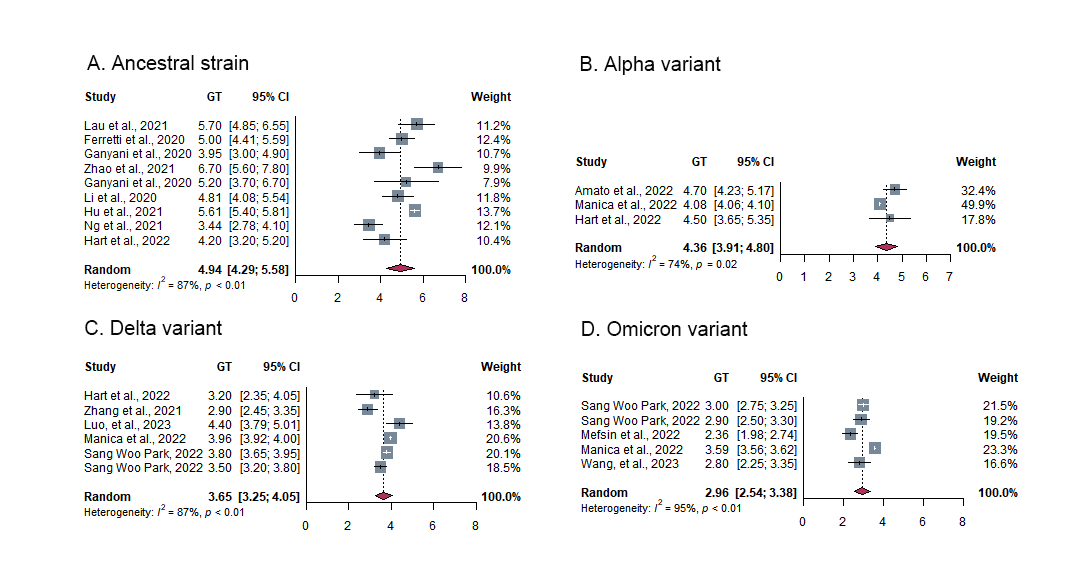

**Figure S3. Forest plot for studies of realized generation time of COVID-19 caused by different SARS-CoV-2 virus lineages.** Twenty-three records were selected and incorporated into meta-analysis. Four records were excluded since they did not provide a central or dispersion tendency estimate, nor could one be inferred from the available summary statistics.

### **Figure S4. Forest plot for studies of intrinsic generation time of COVID-19 caused by different SARS-CoV-2 virus lineages.**

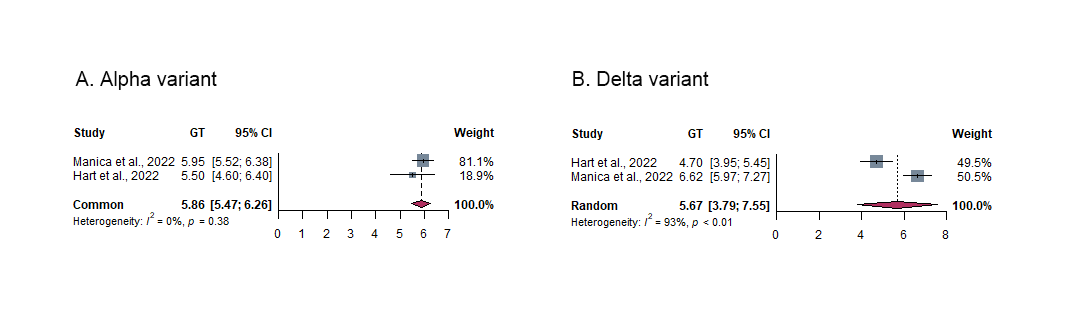

**Figure S4. Forest plot for studies of intrinsic generation time of COVID-19 caused by different SARS-CoV-2 virus lineages.** Four records were selected and included into meta-analysis. The Omicron variant had only one record, thus it was unable to be included.

### Figure S5. Funnel plot for incubation period with 95% confidence interval for included studies in the meta-analysis.

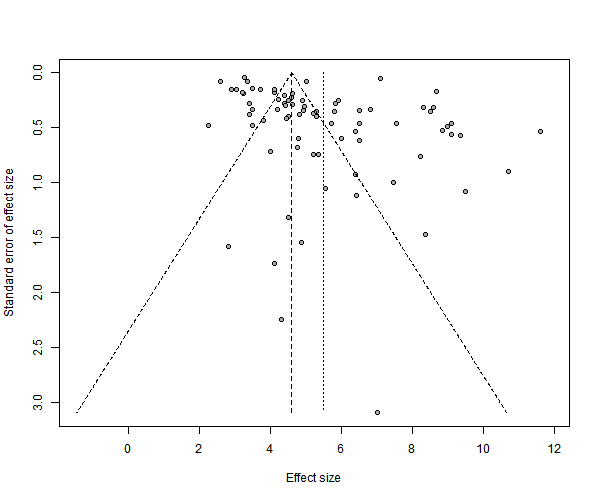

### Figure S6. Funnel plot for serial interval with 95% confidence interval for included studies in the meta-analysis.

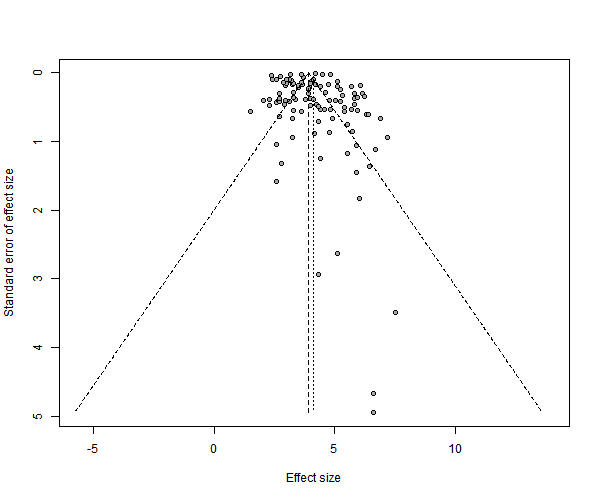

### Figure S7. Funnel plot for generation time with 95% confidence interval for included studies in the meta-analysis.

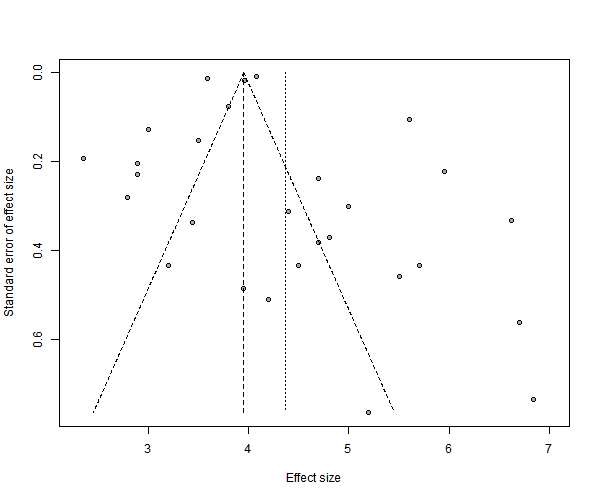
